# Functional near infrared spectroscopy-based dual-stream adaptive language mapping in adults with and without post-stroke aphasia

**DOI:** 10.64898/2026.09.28.26364106

**Authors:** Erin Meier, Veronica Fletcher, Caela Hung, Esprit Ange Andraos, Leanna Ugent, Gengchen Wei, David Lin, Meryem Yücel

## Abstract

In aphasia, variability in lesion location and post-stroke network reorganization complicates efforts to identify the regions that support semantic versus phonological processing. In prior work, researchers have often examined these domains in separate patient cohorts, further limiting direct comparison of their neural substrates within aphasia. To address this gap, we investigated the degree of overlap and specialization within semantic and phonological networks in adults with and without aphasia and examined how neural activity relates to task performance and lesion profiles. Sixteen neurologically healthy adults and 15 participants with aphasia following left hemisphere stroke completed adaptive Semantic Matching and Rhyme Judgment tasks during functional near-infrared spectroscopy (fNIRS) recording. Changes in oxyhemoglobin and deoxyhemoglobin were measured within regions of interest, and activation was related to standardized task performance and damage to left ventral and dorsal language pathways in participants with aphasia. Controls showed largely distinct activation patterns consistent with prior fMRI findings: Semantic Matching preferentially recruited ventral stream regions, particularly left temporal cortex, whereas Rhyme Judgment more strongly recruited dorsal regions, including left inferior frontal and inferior parietal cortex. Participants with aphasia showed greater inter-individual variability and no clear group-level segregation of dorsal and ventral activity. Better Semantic Matching performance was associated with less left ventral stream damage and greater left dorsal activation, whereas better Rhyme Judgment performance was associated with lower right hemisphere ventral and dorsal activation. These findings suggest that post-stroke language performance reflects residual specialization, lesion-dependent constraints, and flexible recruitment of surviving tissue. Overall, fNIRS shows promise as an alternative to fMRI for language mapping in post-stroke aphasia, although replication in larger samples with more extensive cortical coverage is warranted.

## 1. Introduction

The dual-stream model of language processing proposes that a ventral semantic pathway, centered largely in the left temporal cortex, supports the mapping of linguistic representations onto meaning, whereas a predominantly left-lateralized dorsal pathway supports phonological processing and the mapping of phonological representations onto articulatory processes (Hickok & Poeppel, 2004, 2007). A substantial body of converging neuroanatomical and functional evidence underpins this framework, including functional neuroimaging studies in neurologically healthy adults (Binder et al., 2009; Humphreys & Lambon Ralph, 2015; Price, 2012; Vigneau et al., 2006, 2011; Visser et al., 2010) and lesion-symptom mapping studies linking damage to ventral and dorsal stream regions with semantic and phonological impairments, respectively, in individuals with post-stroke aphasia (e.g., Alyahya et al., 2020; Butler et al., 2014; Halai et al., 2017, 2018; Henseler et al., 2014; Lacey et al., 2017; Mirman, Chen, et al., 2015; Mirman, Zhang, et al., 2015; Schumacher et al., 2019; Schwartz et al., 2009, 2012; Walker et al., 2011).

Recent meta-analytic evidence indicates that semantic and phonological tasks recruit partially overlapping but reliably dissociable cortical networks, further supporting the functional organization proposed by dual-stream models (Hodgson et al., 2021; Turker et al., 2023).

This framework provides a useful basis for investigating how semantic and phonological functions are reorganized following stroke. Despite damage to canonical language regions, people with aphasia (PWA) retain considerable capacity for language gains following treatments targeting semantic and phonological processes (Brady et al., 2016; Efstratiadou et al., 2018; Python et al., 2025; Quique et al., 2019). Such recovery may depend not only on surviving components of the system typically supporting a given linguistic function, but also on functional redundancy within the broader language network or domain-general systems (Billot & Kiran, 2024; Hartwigsen & Saur, 2019; Stefaniak et al., 2020; Turkeltaub, 2019). Indeed, at the chronic stroke stage, functional neuroimaging studies have implicated combinations of residual left hemisphere regions in both streams, their right hemisphere homologues, and regions not traditionally associated with linguistic processing during language production and comprehension tasks (LaCroix et al., 2021; Stefaniak et al., 2021; Turkeltaub et al., 2011; Wilson & Schneck, 2021). Pillay et al. (2022) demonstrated that in individuals with chronic left hemisphere stroke and phonological impairment, successful reading was associated with greater recruitment of the residual semantic network rather than the phonological network, suggesting that surviving circuitry associated with one linguistic subsystem may sometimes support performance when another is compromised.

Determining the extent of semantic and phonological cross-domain recruitment is challenging, however, because most functional neuroimaging studies of aphasia have employed a single language paradigm. This approach makes it difficult to determine whether activity observed during semantic or phonological processing reflects specialization for that domain or more general recruitment across language tasks (Diedrichs et al., 2026). Few studies have directly examined both processes within the same individuals. Fernandez et al. (2004), for example, longitudinally examined semantic and phonological activation maps in an individual with aphasia and observed both task-related differentiation and recruitment of a broader bilateral network during recovery. Rochon et al. (2010) similarly compared picture-based semantic and rhyme judgments in four PWA, and although the tasks elicited partially distinct patterns of activity, activation extended beyond canonical semantic and phonological regions, with some regions recruited across both tasks and considerable variability among patients. More recently, Casilio et al. (2024) demonstrated partly overlapping recruitment across written rhyme judgment and semantic matching tasks in a longitudinal case participant with aphasia with word deafness. Hartwigsen et al. (2020) examined semantic and phonological word decisions in 12 individuals with chronic left temporoparietal stroke and discovered at baseline, both tasks recruited an extensive, largely overlapping bilateral network, including inferior frontal, insular, supplementary motor, and other frontal regions. Yet, perturbation of the left inferior frontal gyrus (IFG) revealed a functional-anatomical dissociation: disruption of the posterior IFG selectively impaired phonological decisions, whereas disruption of the anterior IFG preferentially affected semantic decisions. Moreover, stimulation-induced delays in phonological processing speed over the posterior IFG were associated with increased recruitment of the contralesional right supramarginal gyrus (SMG), providing evidence for both persistent functional specificity and flexible compensatory recruitment within the damaged language system.

Collectively, this literature suggests that semantic and phonological processing after stroke may exhibit relative rather than absolute functional specialization. Regions classically associated with each linguistic system continue to demonstrate domain-specific contributions, yet considerable overlap and cross-domain recruitment are also observed, with their extent likely shaped by lesion distribution and the availability of surviving network resources. Inter-individual variability further complicates conclusions drawn by comparing separate groups performing different paradigms. Assessing semantic and phonological processing within the same individual (in this case PWA) therefore provides an important means of distinguishing regions that retain relatively domain-specific functions from those that participate more flexibly across linguistic processes following stroke.

The current study investigated the degree of overlap and specialization within semantic and phonological networks in adults with and without aphasia using the Adaptive Language Mapping (ALM) paradigms developed by Wilson and colleagues (Wilson et al., 2018; Yen et al., 2019) and an emergent imaging technology for investigating neurorecovery, functional near-infrared spectroscopy (fNIRS). A critical challenge in interpreting functional neuroimaging studies of aphasia is that differences in activation may reflect differences in task performance or difficulty rather than changes in language organization itself (Wilson & Schneck, 2021). The ALM paradigms were designed to mitigate this problem by dynamically adjusting stimulus complexity and presentation rate according to individual performance. In fMRI, the adaptive semantic paradigm has demonstrated feasibility and reliable language mapping in PWA, while the complementary adaptive rhyme judgment paradigm reliably identifies regions implicated in phonological processing in healthy adults (Wilson et al., 2018; Yen et al., 2019). Thus, the two paradigms provide a method for probing distinct linguistic processes while more closely equating performance demands across individuals with widely varying language abilities. Here, we measured neural responses to these paradigms using fNIRS, a noninvasive optical imaging approach that estimates cortical activity from task-related changes in oxygenated and deoxygenated hemoglobin. Compared with fMRI, fNIRS is portable, more accessible, more tolerant of modest movement, and free from many MRI contraindications, making it particularly attractive for functional imaging in clinical populations (Butler et al., 2020; Pinti et al., 2020; Quaresima & Ferrari, 2019).

Thus, the novelty of the present study lies in integrating matched semantic and phonological paradigms within the same individuals with post-stroke aphasia, while simultaneously relating task-specific functional recruitment to behavioral performance and structural damage within dorsal and ventral language pathways. This design allows us to distinguish residual functional specialization from broader cross-domain recruitment in a way that cannot be readily resolved when these components are examined separately. Based on findings from reviews of fMRI studies in healthy adults (Diedrichs et al., 2026; Hodgson et al., 2021; Noonan et al., 2013; Turker et al., 2023) and work by Wilson and colleagues (Wilson et al., 2018; Yen et al., 2019), we predicted neurologically healthy adults would demonstrate left-lateralized activation for both tasks with functional dissociation of key dorsal (i.e., left posterior IFG, left SMG) and ventral (i.e., left anterior IFG, middle and posterior inferior temporal gyrus) stream hubs for phonological and semantic tasks, respectively. Based on the few dual-paradigm studies in PWA and broader language meta-analyses in chronic aphasia (LaCroix et al., 2021; Stefaniak et al., 2021; Turkeltaub et al., 2011; Wilson & Schneck, 2021), we expected as a group, PWA would demonstrate greater right hemisphere engagement and less functional specialization than controls, indicated by some regions activated for both tasks. However, we also predicted that better task performance within PWA would be associated with greater recruitment of residual tissue within left dorsal and ventral stream hubs for the respective phonological and semantic paradigms. We additionally expected that greater damage to these hubs would be associated with either higher engagement of their right hemisphere homologues or opposite-stream regions, consistent with theory regarding network degeneracy (Stefaniak et al., 2020).

## 2. Materials and Methods

### 2.1. Participants

Participants were recruited via flyers posted on social media and approved locations and through established pipelines within The Aphasia Network Laboratory at Northeastern University. Inclusion criteria for both groups were normal or corrected-to-normal vision and hearing; current or pre-stroke proficiency in English; and no neurological conditions known to affect cognitive-linguistic skills (besides stroke in the patient group). Exclusion criteria were major psychiatric diagnoses (e.g., schizophrenia), any major medical conditions (e.g., cancer) that would preclude study participation, and for controls, presence of cognitive deficits based on the Mini Mental State Examination (MMSE) education-adjusted cutoff (Folstein et al., 1983).

Fifteen individuals with a history of chronic left hemisphere stroke and 16 neurologically healthy controls were enrolled and completed study procedures. Participants with a history of stroke were administered the Western Aphasia Battery-Revised (WAB-R; Kertesz, 2007), from which the Aphasia Quotient (AQ) was derived to determine presence of aphasia. Four individuals scored above the WAB-R AQ cutoff but had a history of an aphasia diagnosis and endorsed lingering difficulty with language production that differed from their premorbid levels. Subject-level demographic, testing, and stroke data are reported in Table 1.

**Table 1.**
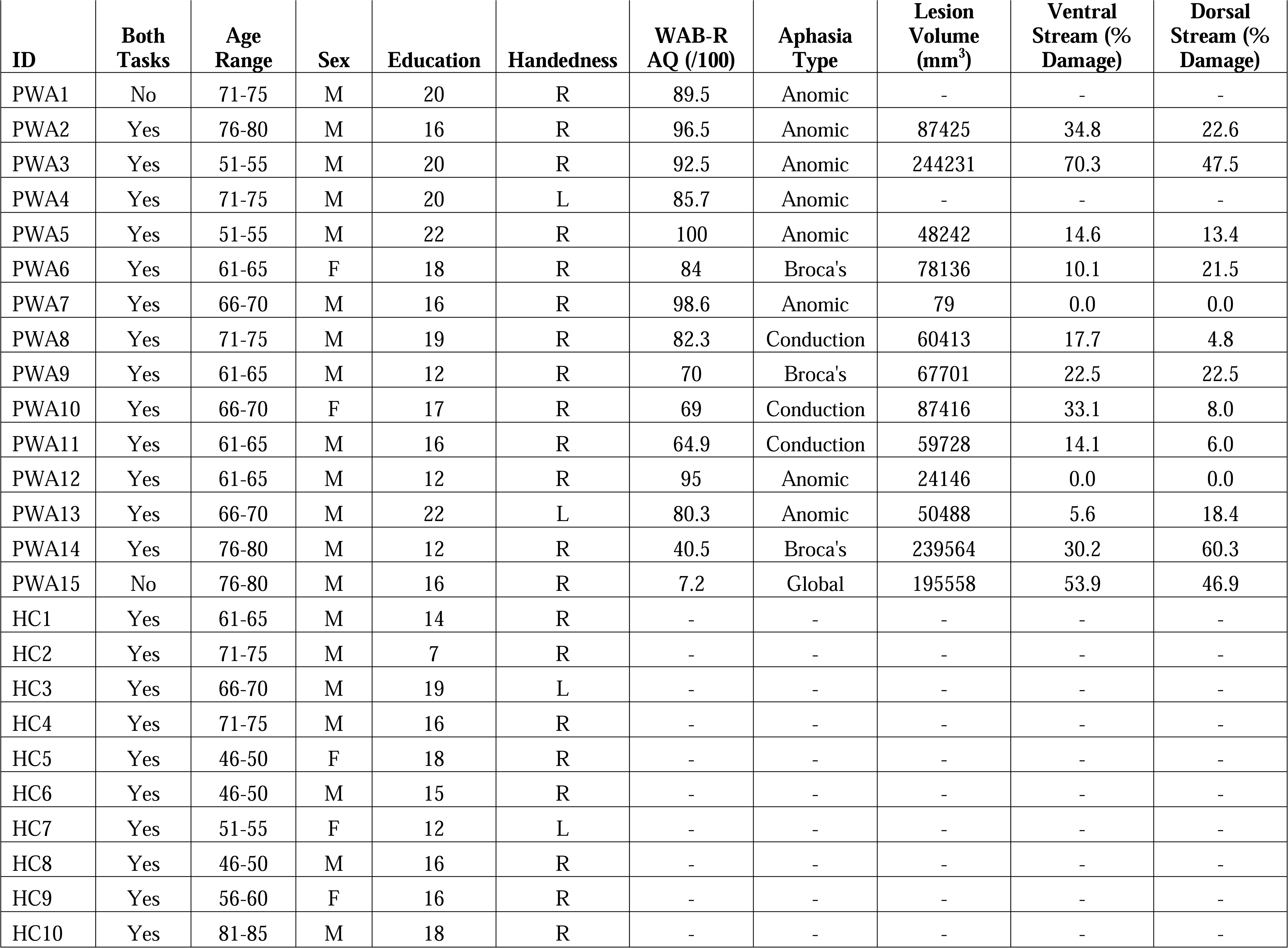
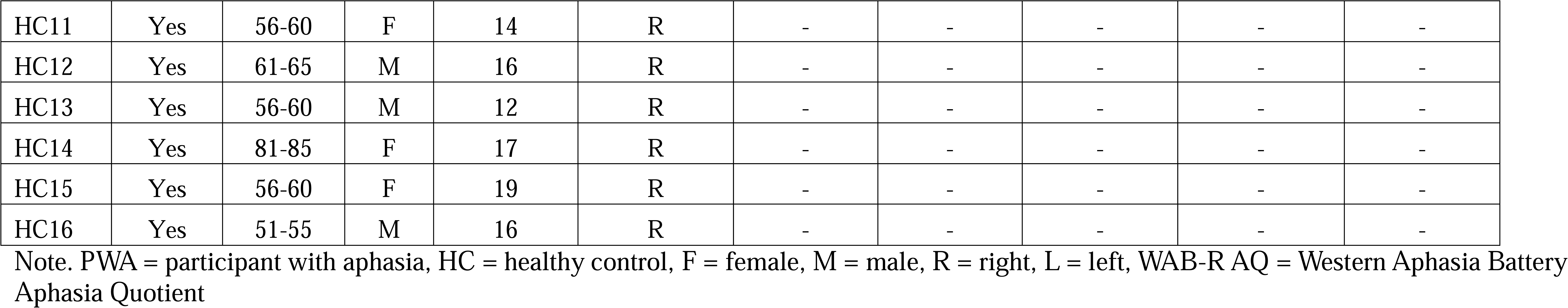
Sample Demographic and Stroke Characteristics.

Study procedures were carried out in accordance with the Declaration of Helsinki and the Northeastern University Department of Human Research Institutional Review Board (protocol #21-06-11). All participants provided their written informed consent prior to completing any study activities.

### 2.2. Adaptive Language Mapping Tasks

Participants completed one run of the written Semantic Matching (Wilson et al., 2018) and written Rhyme Judgment (Yen et al., 2019) ALM tasks, presented using MATLAB and Psychtoolbox-3 (Brainard, 1997; Kleiner et al., 2007; Pelli, 1997). Both paradigms followed an AB block design with alternating 20 s blocks of experimental and control trials. Each task contained 10 blocks of each condition. In the Semantic Matching experimental condition trials, participants decided whether two written words (e.g., *cat-mouse*, *boy-floor*) were related in meaning. In the Rhyme Judgment experimental condition trials, participants judged whether two written nonwords (e.g., *pachine-washeen*, *lecovery-picovany*) rhymed. In both tasks, the control condition was a non-linguistic perceptual decision task during which participants judged whether two faux letter strings (e.g., ЖЖ□-ЖЖ□, Œ□□δ□□□□-Œ□□δ□□Ə□) were an exact match. For both tasks and conditions, participants were instructed to push a button with their left hand for a match and to do nothing for a mismatch. Difficulty varied by level (via a two-down/one-up adaptive staircase with weighted step sizes) and trial presentation rate. Thus, while the block duration was held constant, the number and difficulty of trials varied across blocks within and between participants. More details regarding the paradigms can be found in the publications by Wilson and colleagues (Wilson et al., 2018; Yen et al., 2019). The ALM paradigms are freely available at https://aphasialab.org/alm/.

To characterize global behavioral performance on the fNIRS tasks within the group of PWA, we created a composite performance score for every participant in the aphasia group. First, using the z-score formula (x□*-m/s*), a standardized score for combined accuracy and response time (RT) was generated for each individual with aphasia based on control performance, where x□ was average accuracy/RT for an individual with aphasia, and *m* and *s* corresponded, respectively, to the control sample’s mean accuracy/RT and standard deviation for a given paradigm. This procedure resulted in standardized accuracy and RT scores where positive values were interpreted as better accuracy but worse response time performance. Thus, to create a single composite score for Semantic Matching and Rhyme Judgment for each participant with aphasia, the sign for the RT z-score was flipped and the two task scores were averaged. These composites were used to address relationships between task performance and brain metrics within the PWA group.

### 2.3. fNIRS Data Acquisition

Like fMRI, fNIRS relies on neurovascular coupling to provide indirect estimates of neural activity associated with changes in cortical hemodynamics. Source optodes placed on the scalp emit low-intensity near-infrared light, which propagates through extracerebral tissues and penetrates the underlying cortical tissue. Oxyhemoglobin (HbO) and deoxyhemoglobin (HbR) differentially absorb near-infrared light within the approximate 650-900 nm range, allowing task-related changes in their concentrations to be estimated from changes in detected light intensity. A portion of the diffusely scattered light returns to the scalp surface, where it is measured by photodetectors. Changes in detected light intensity are then converted into relative changes in HbO and HbR concentrations. Each source-detector pair forms a measurement channel that is sensitive to tissue along its photon propagation path, with the midpoint between the source and detector commonly used to approximate the channel’s cortical location. The canonical fNIRS hemodynamic response is characterized by an increase in HbO accompanied by a smaller decrease in HbR, emerging several seconds after stimulus onset and typically reaching its peak during the subsequent 5-10 s, although the timing and shape of the response vary with task and stimulus duration.

The fNIRS data were acquired with two daisy-chained 8×8 NIRx NIRSport2 continuous wave systems (https://nirx.net/nirsport) at a sampling rate of 10.2 Hz. The connected systems resulted in a total of 16 dual-tipped LED sources that emitted near-infrared light at two wavelengths (760 and 850 nm) and 16 dual-tipped silicon photodiode detectors. Paired sources and detectors were arranged over perisylvian regions and spaced ≤ 30 mm apart to form a symmetrical array of 22 long-distance measurement channels within each cerebral hemisphere. One detector (D16) was sacrificed to form eight, 8mm short-distance channels with each paired source (i.e., S01, S02, S06, S08, S09, S10, S14, and S16). Short distance channels were used to measure superficial physiological signals (e.g., blood flow in the meningeal arteries), which were regressed from the long-distance channel data.

The fNIRS probe was designed to measure activity from cortical regions traditionally associated with domain-general non-linguistic processes, including the bilateral prefrontal cortices, and areas within the bilateral dorsal and ventral language pathways. Thus, bilateral regions covered by the probe included portions of the frontal pole, rostrolateral prefrontal cortex (RLPFC), and dorsolateral prefrontal cortex (DLPFC) that included the anterior middle and superior frontal gyri (MFG and SFG, respectively); the inferior frontal gyrus, pars triangularis (IFGtri) and pars opercularis (IFGop); the mid to posterior superior (STG), middle (MTG), and inferior (ITG) temporal gyri; and the supramarginal gyrus (SMG) and angular gyrus (AG) in the parietal lobe. The MNI coordinates corresponding to the midpoint of each source-detector pair and the associated Automated Anatomical Labeling (AAL) Atlas (Tzourio-Mazoyer et al., 2002) regional label were extracted from AtlasViewer (Aasted et al., 2015). Channels were subsequently grouped into regions of interest (ROIs) based on the approximate MNI coordinate locations (Gilmore et al., 2021; Li et al., 2020). Figure 1 shows the probe design, measurement sensitivity profile, and channel-to-ROI assignment. Supplemental Table 1 reports the MNI coordinate of the center of each channel and assignment of ROIs to the ventral and dorsal language pathways. Note that bilateral SFG was not assigned to either language pathway given its channels’ proximity to the domain-general, multiple-demand system (Duncan, 2010; Fedorenko et al., 2011). Both parts of IFG were assigned to both language pathways given somewhat conflicting evidence regarding their specific contributions to semantic versus phonological processing (Hodgson et al., 2021; Liakakis et al., 2011; Vigneau et al., 2006).

**Figure 1.**
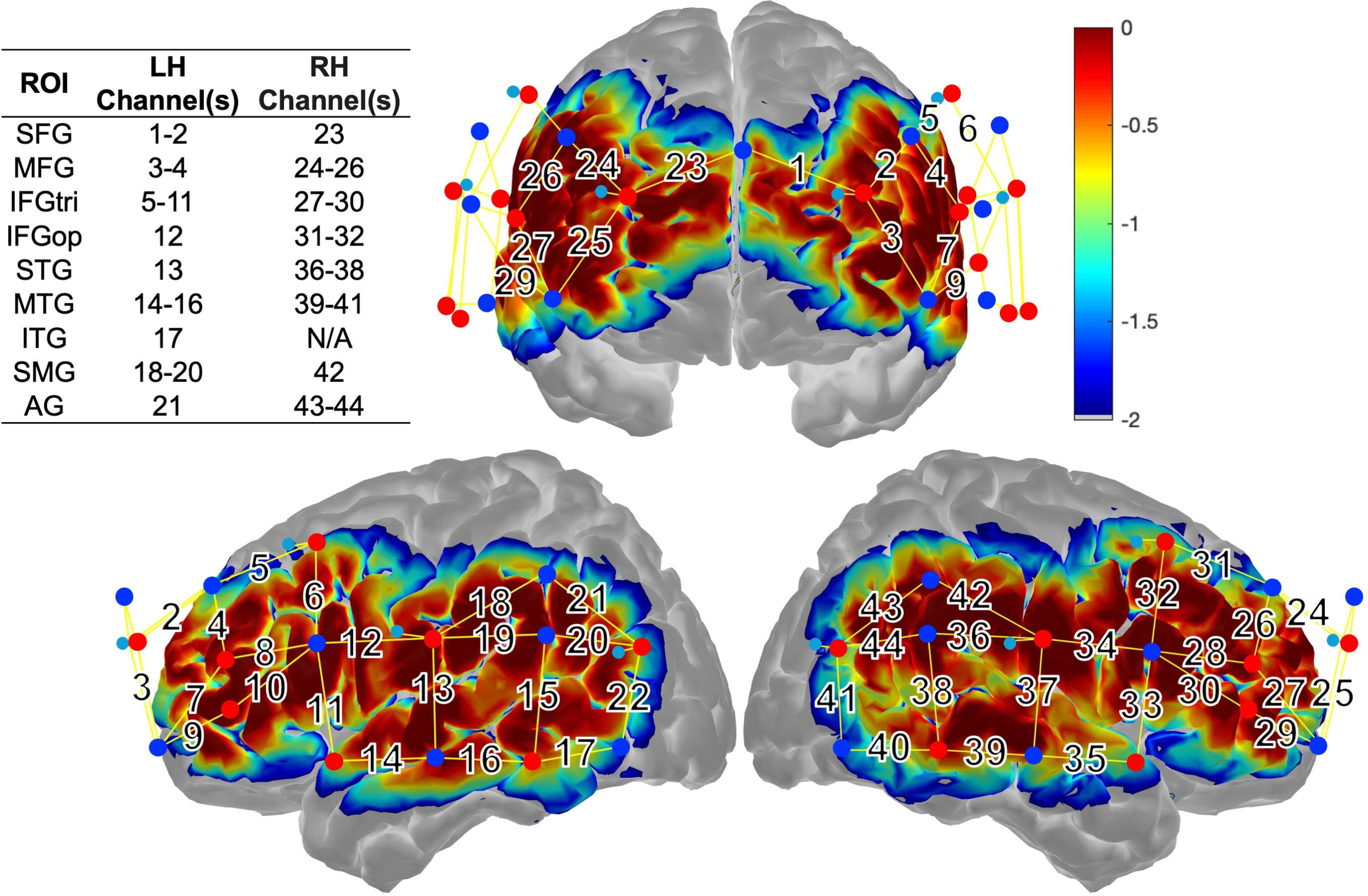
fNIRS probe design and measurement sensitivity profile. Sixteen light-emitting sources (denoted by red circles) and 16 photodiode detectors (denoted by royal blue circles) formed 44 long-distance measurement channels (numbered and denoted by solid yellow lines) over bilateral perisylvian cortex. Short-distance detectors (denoted by small, light blue circles) placed in close proximity to two anterior and two posterior sources per hemisphere formed eight short-distance channels. Areas of higher measurement sensitivity are denoted by warm colors; cool colors reflects areas of lower sensitivity. Channels that comprise regions of interest (ROIs) are denoted in the upper left inset. AG = angular gyrus, IFGtri/op = inferior frontal gyrus, pars triangularis/opercularis, ITG = inferior temporal gyrus, LH = left hemisphere, MFG = middle frontal gyrus, MTG = middle temporal gyrus, RH = right hemisphere, SFG = superior frontal gyrus, SMG = supramarginal gyrus, STG = superior temporal gyrus.

Prior to fNIRS data acquisition, participants underwent task training and practice as described in Wilson et al. (2018) and Yen et al. (2019). Study team members measured each participant’s head circumference, the nasion-to-inion distance, and the distance between their left and right pre-auricular points to select the correctly sized cap and ensure proper placement of the cap based on the location of cap fiducial markers (e.g., Cz) relative to the participants’ head anatomy. After the cap was placed, the signal optimization procedure in Aurora (https://nirx.net/software) was performed to evaluate source signal levels, the amount of dark noise (reflecting interference with light returning to the detectors), and signal-to-noise ratio (SNR) for each sensor. Suboptimal readings (i.e., “critical” flag in Aurora) were improved by parting the hair, loosening the sensor wires, and/or replacing low-tension with high-tension spring-loaded optode tops to ensure good coupling of the sensors against the scalp as well as placing an opaque shower cap over the optode cap to block external light. The fNIRS data were also recorded in Aurora, and the quality of incoming signals were monitored throughout acquisition. Optode adjustments and re-running the signal optimization procedure were done as needed between tasks.

### 2.4. Structural Imaging and Lesion Approach

Structural images were acquired to account for lesioned tissue in the fNIRS analysis. For a subset of the participants with aphasia (*n* = 7), lesions were traced in MRIcron (Rorden & Brett, 2000) on a high-resolution T1-weighted MPRAGE scan. For other PWA (*n* = 6) with MRI contraindications (e.g., implanted medical devices) or who declined undergoing research-grade MRI scanning, lesions were traced on clinical CT or MRI scans. Traced lesion maps and the associated structural scan were warped to a standard older adult template using the Clinical Toolbox (Rorden et al., 2012) in SPM12 (https://www.fil.ion.ucl.ac.uk/spm/software/spm12/).

Bespoke MATLAB scripts based on routines from MarsBaR (Brett et al., 2002) were used to calculate overall lesion volume and the percentage of damaged tissue within ROIs contributing to the dorsal and ventral pathways (see Supplemental Table 1). Following the approach by (Gilmore et al., 2021), lesions were masked out from the fNIRS data by manual removal of data from channels over lesioned tissue. See Figure 2 for a depiction of overlaid lesions across PWA.

**Figure 2.**
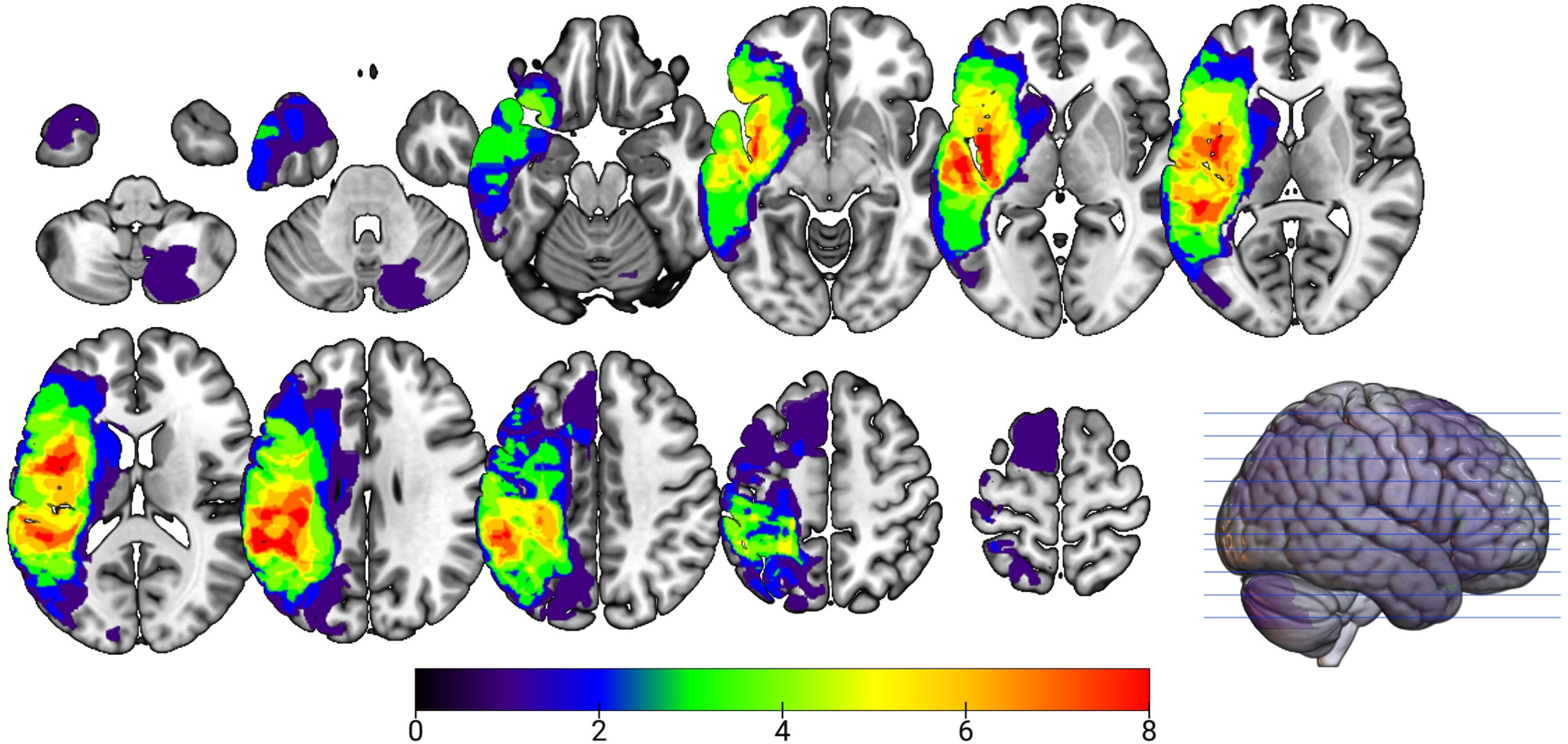
Lesion overlay in participants with aphasia (*n* = 13). Areas of greatest overlap are reflected by warmer colors whereas regions of lower overlap are shown in cooler colors. Structural images were unavailable for two participants with aphasia (PWA1 and PWA4).

Of note, structural images could not be obtained for PWA1 and PWA4. Because only very general descriptions regarding lesion location were available (i.e., affected hemisphere and anterior versus posterior focus), we took a conservative approach to the fNIRS analysis for these two PWA by removing all left hemisphere channels. We believe this approach was best to minimize confounds in this small pilot sample given that near-infrared light diffuses across all tissue types before re-emerging to the head surface, so there is no inherent masking of the lesion contained within the processed fNIRS time series.

### 2.5. fNIRS Data Analysis

Data were preprocessed in Homer3 (Huppert et al., 2009) following a standard pipeline for fNIRS task data. First, channels with SNR levels below 5.0 were pruned, followed by the conversion of raw signals to changes in optical density (OD). Next, motion artifacts were corrected using a combination of spline interpolation and Savitzky-Golay filtering (Jahani et al., 2018) and visually inspected by study team members. Any residual spikes within the OD data 200 ms surrounding the stimulus onset were flagged in an artifact detection step, and the stimulus markers surrounding these uncorrected artifacts were rejected, resulting in the removal of the corresponding block. The OD data were then high-pass filtered (at 0.01 Hz) to remove low-frequency noise (e.g., slow signal drift) and low-passed filtered (at 0.5 Hz) to remove high-frequency noise (e.g., cardiac pulsations). Next, changes in OD were converted into relative changes in oxyhemoglobin (HbO) and deoxyhemoglobin (HbR) concentrations via the modified Beer-Lambert Law (Kocsis et al., 2006). No partial path length correction was applied. The hemodynamic response function (HRF) was modeled using a sequence of Gaussian basis functions with a 1-s standard deviation and 1-s temporal spacing, spanning 2 s before to 25 s after each block onset (Diamond et al., 2006). The GLM also incorporated a short-separation regression using the short-distance data most highly correlated with the long-distance channel data (Gagnon et al., 2012; Yücel et al., 2015). Finally, changes in HbO and HbR concentrations for the contrast of the experimental > control trials were calculated using an ordinary least squares general linear model (GLM).

Across healthy controls, the average percentage of pruned long-distance channels was 1.47% (SD = 3.76%) for Semantic Matching and 2.70% (SD = 6.95%) for Rhyme Judgment. Thirteen (out of 16) controls had no pruned long-distance channels for either task. Two healthy controls with thick, dark hair had the most pruned channels for both tasks, with five and nine pruned channels for Semantic Matching and Rhyme Judgment, respectively, in both individuals’ datasets. Despite the higher number of pruned channels compared to other controls, data from both participants were retained for analysis as they each had greater than 75% good-quality channels from both tasks. There were no pruned long-distance channels due to residual artifacts for either task in the PWA group. Supplemental Table 1 lists pruned and lesioned/removed channels in healthy control and PWA datasets, respectively.

### 2.6. Statistical Analyses

Post-processing and statistical analysis of the preprocessed fNIRS time series was done in R (R Core Team, 2024). First, to evaluate activation patterns, HbO and HbR concentration data from each task were downsampled to 1 s epochs by averaging across the datapoints collected within each time window across the time series (-2 to 25 s). After the removal of lesioned channels from the PWAs’ datasets (see section 2.4), the downsampled data were averaged across channels corresponding to each ROI (per Figure 1 and Supplemental Table 1).

To assess which regions were activated by each task contrast within each group, we first plotted the ROI-level HbO and HbR concentrations, determining which regions demonstrated a canonical hemodynamic response to the tasks (i.e., an increase in HbO and corresponding decrease in HbR concentrations over time). To identify significant within-group task-related brain activity, we used the *nlme* package (Pinheiro et al., 2023; Pinheiro & Bates, 2000) to fit a series of linear mixed-effects models (LMEMs) separately for each ROI and hemoglobin species over the primary response window (5-20 s). Time was modeled as a categorical fixed effect, with participant included as a random intercept and an AR(1) structure used to model residual autocorrelation across time. Model-level *p*-values were corrected for multiple comparisons across ROIs using a false discovery rate (FDR) threshold of *q* < 0.05. Estimated marginal means were subsequently used within *emmeans* (Lenth, 2024) to identify individual time points at which HbO was significantly greater than zero and HbR was significantly less than zero. Both the statistical significance and morphology of the HbO and HbR response curves were considered when determining presence of significant brain activity within each group; however, greater weight was placed on the canonical HbO response because of its higher SNR (Pan et al., 2019; Strangman et al., 2002).

To assess between-group differences in brain activity, a similar series of LMEMs were fit with Group, Time, and Group x Time interaction modeled as fixed effects, with Group and Time modeled categorically, and the reference level set to the PWA group. Participant was included as a random intercept, and an AR(1) structure was also used to model residual autocorrelation across time. Similar to the within-group analytic approach, model-level *p*-values were corrected for multiple comparisons using an FDR threshold of *q* < 0.05. For models in which the omnibus Group x Time effect survived FDR correction, estimated marginal means were used to identify individual time points at which PWA and controls differed; these pairwise comparisons were two-sided. The primary goals of this analysis were to assess between-group differences in task-evoked responses and to determine whether the temporal profile of the response differed between PWA and controls. Accordingly, although main effects of time are reported, interpretation focuses on effects of Group and Group x Time.

To examine between-group differences in task performance, we first used the *afex* package (Singmann et al., 2025) to conduct a series of 2 (Group) x 2 (Condition) mixed ANOVAs examining percent accuracy, mean RT, and mean trial difficulty separately for each task. To examine brain-behavior relationships within PWA, channel-level HbO values during the expected peak activation window for each block (5-20 s) were averaged across time and across channels within each language stream by hemisphere. For each task, Spearman correlations were then used to examine associations between the standardized composite behavioral measure and six neural measures: lesion burden within the left ventral and dorsal streams, and activation within the ventral and dorsal streams of the left and right hemispheres.

### 2.7. Data Availability Statement

The datasets analyzed for this study are available on Open Science Framework at https://osf.io/zk4j5/overview.

## 3. Results

Participants in the control and stroke groups did not differ in age (*t*_26.2_ =-1.58, 95% CI [-12.43, 1.64], *p* = 0.127), sex (OR = 3.73, 95% CI [0.52, 45.56], *p* = 0.220), handedness (OR = 0.93, 95% CI [0.06, 14.61], *p* = 1.00), or years of education (*t*_28.2_ =-1.62, 95% CI [-4.27, 0.49], *p* = 0.116). All healthy controls completed both ALM paradigms while 13 of the 15 PWA completed both tasks. PWA1 and PWA15 did not complete the Rhyme Judgment task due to below-chance performance even after repeated training attempts during the task training phase.

### 3.1. Adaptive Language Mapping Activation Patterns

#### 3.1.1. Semantic Matching

Figure 3 shows changes in HbO and HbR for Semantic Matching in left and right hemisphere ROIs over the modeled block period (-2 to 25 s). Table 2 reports significant within-group omnibus model effects for PWA and controls, while Supplemental Tables 2 and 3 report the full within-group LMEM outputs—including time point findings—for Semantic Matching for controls and PWA, respectively.

**Figure 3.**
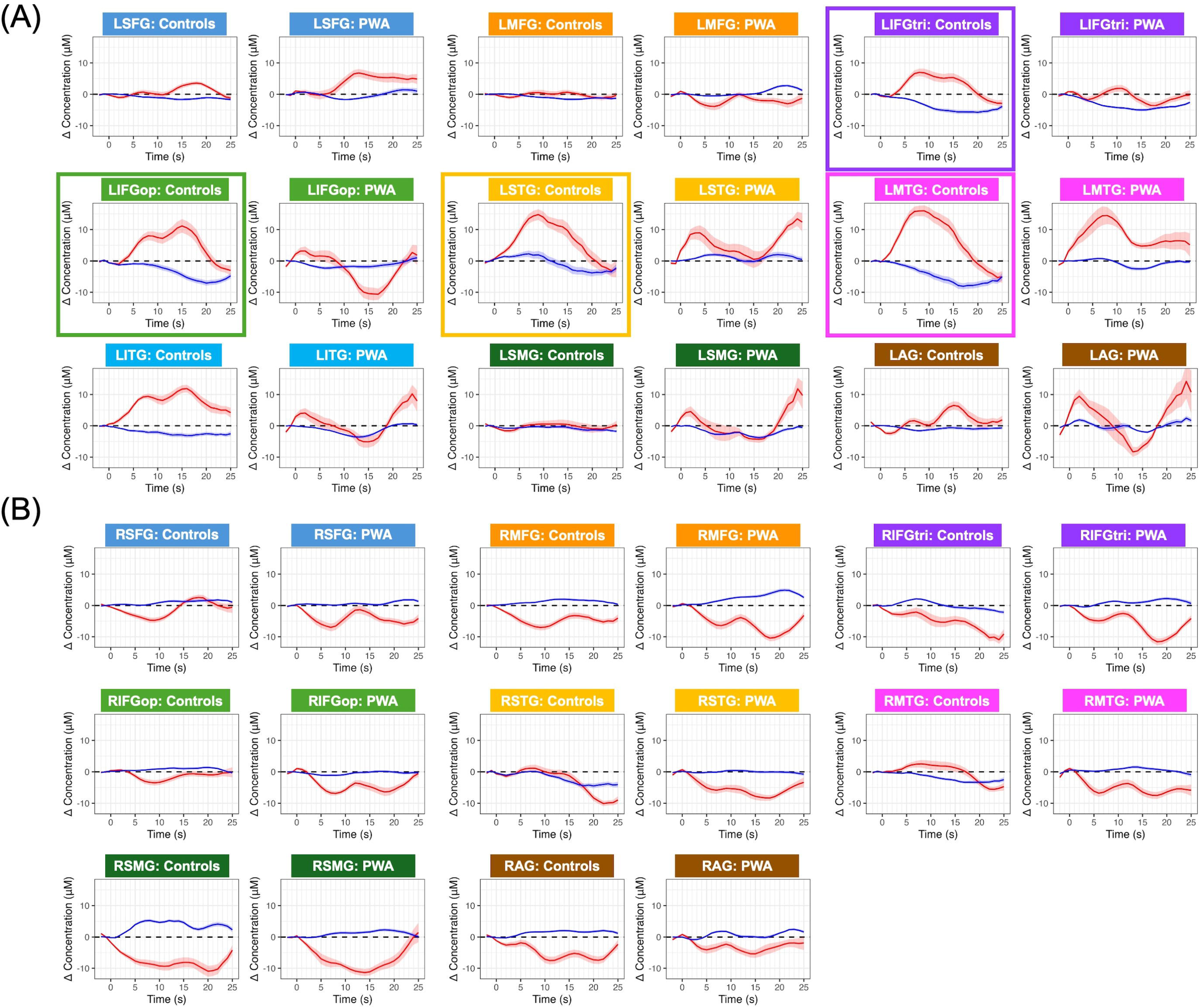
Changes in oxyhemoglobin (in red) and deoxyhemoglobin (in blue) for left hemisphere (A) and right hemisphere (B) regions of interest during Semantic Matching. For each region of interest (ROI), the left-side plot shows the group-averaged hemodynamic GLM output for controls, and the right-side plot shows the group-averaged hemodynamic GLM output for participants with aphasia. Solid color borders reflect ROIs for which a canonical and significant oxyhemoglobin time course was observed. Shaded ribbons represent within-group standard error around the mean time series. AG = angular gyrus, IFGtri/op = inferior frontal gyrus, pars triangularis/opercularis, ITG = inferior temporal gyrus, LH = left hemisphere, MFG = middle frontal gyrus, MTG = middle temporal gyrus, RH = right hemisphere, SFG = superior frontal gyrus, SMG = supramarginal gyrus, STG = superior temporal gyrus.

**Table 2.** Significant Within-Group Time Effects During Semantic Matching.

| Group | Hemoglobin | Region | N | F | df | p | q (FDR) |
| --- | --- | --- | --- | --- | --- | --- | --- |
| Controls | HbO |  |  |  |  |  |  |
|  |  | LIFGtri | 16 | 3.18 | 15, 225 | < .001 | .002 |
|  |  | LIFGop | 16 | 2.52 | 15, 225 | .002 | .010 |
|  |  | LSTG | 16 | 2.13 | 15, 225 | .010 | .033 |
|  |  | LMTG | 16 | 2.89 | 15, 225 | < .001 | .003 |
|  |  | RSFG | 16 | 1.93 | 15, 225 | .022 | .061 |
|  |  | RSTG | 16 | 2.18 | 15, 225 | .008 | .033 |
|  | HbR |  |  |  |  |  |  |
|  |  | LIFGtri | 16 | 3.12 | 15, 225 | < .001 | .002 |
|  |  | RIFGtri | 16 | 1.74 | 15, 225 | .045 | .240 |
|  |  | RSTG | 16 | 1.91 | 15, 225 | .023 | .199 |
| PWA | HbO |  |  |  |  |  |  |
|  |  | RMFG | 15 | 2.19 | 15, 210 | .008 | .066 |
|  |  | RIFGtri | 15 | 2.64 | 15, 210 | .001 | .019 |
|  | HbR |  |  |  |  |  |  |
|  |  | LSFG | 15 | 1.85 | 15, 210 | .029 | .100 |
|  |  | LMFG | 15 | 4.12 | 15, 210 | < .001 | < .001 |
|  |  | LIFGtri | 13 | 2.1 | 15, 180 | .012 | .049 |
|  |  | LITG | 12 | 2.17 | 15, 165 | .009 | .049 |
|  |  | RMFG | 15 | 3.11 | 15, 210 | < .001 | .001 |

Note. Only omnibus time effects with uncorrected $p < 0.05$ or FDR-corrected $q < 0.05$ are shown. Full model outputs are reported in Supplement Table 2 for controls and Supplemental Table 3 for participants with aphasia (PWA). AG = angular gyrus, HbO = oxyhemoglobin, HbR = deoxyhemoglobin, IFGtri/op = inferior frontal gyrus, pars triangularis/opercularis, ITG = inferior temporal gyrus, L = left, MFG = middle frontal gyrus, MTG = middle temporal gyrus, R = right, SFG = superior frontal gyrus, SMG = supramarginal gyrus, STG = superior temporal gyrus.

**Table 3.**
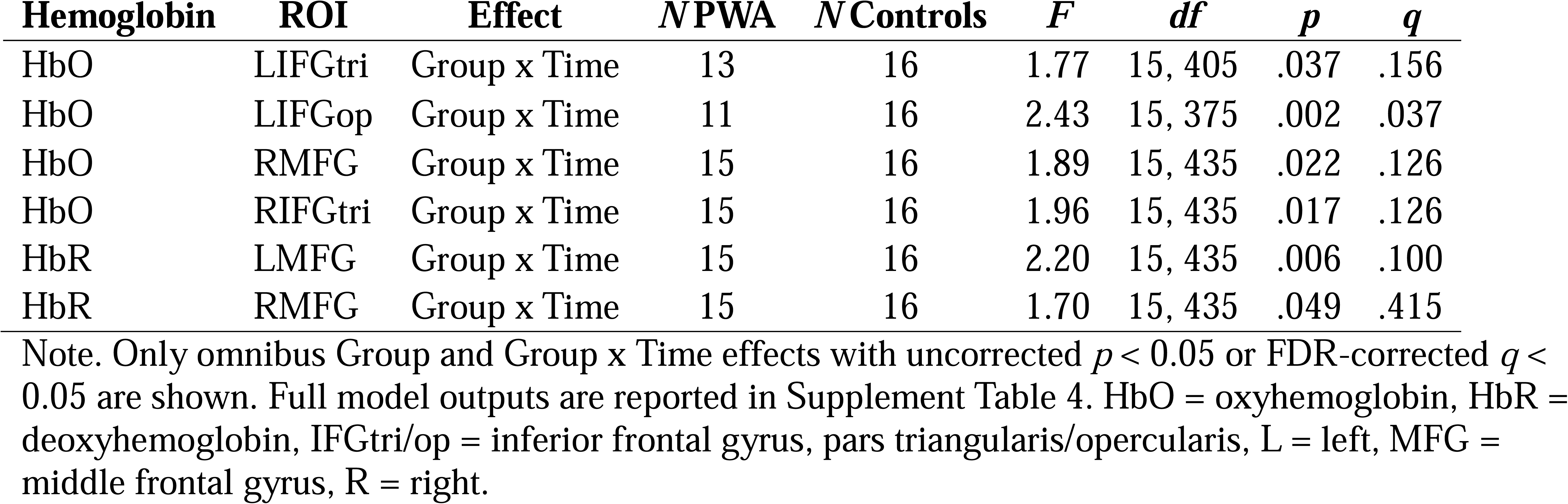
Significant Effects of Group and Group x Time During Semantic Matching.

Based on visual inspection of the time series plots and confirmed with the LMEMs, healthy controls exhibited a significant increase in HbO from 7-10 s at an uncorrected threshold, with a corresponding significant decrease in HbR within left IFGtri from 9-20 s during Semantic Matching (Figure 3A, Table 2). Canonical response curves and significant increases in HbO without significant decreases in HbR were observed in left IFGop from 13-17 s, left STG from 6-14 s, and left MTG from 5-15 s. As reported in Table 2, control LMEMs contained significant main effects of time for one or both hemoglobin species for right SFG, IFGtri, and STG, but response morphology reflected deactivation, rather than activation (see Figure 3B). As illustrated in Figure 3A, controls also exhibited expected response curves for both hemoglobin species within left SFG, left ITG—and to a lesser extent—left AG, but omnibus LMEM effects were not significant for these regions (Supplemental Table 2). At a group level, PWA exhibited somewhat canonical hemodynamic responses within left IFGtri and left MTG, but only the omnibus test of time for left IFGtri HbR was statistically significant, with effects across the entire time window (5-20 s) (see Table 2 and Supplemental Table 3). Other significant model effects reported in Table 2 reflect response curves consistent with deactivation for PWA. No other left hemisphere ROIs were recruited at a group level across PWA nor were any right hemisphere ROIs significantly activated by either group.

Significant findings from the between-group LMEMs are reported in Table 3, and the full LMEM model output is reported in Supplemental Table 4. A significant Group x Time interaction was observed for HbO in left IFGtri, left IFGop, right MFG, and right IFGtri as well as HbR in bilateral MFG. However, only the effect for LIFGop HbO survived model-level correction for multiple comparisons; negative estimates in the latter portion of the time window (13-18 s) (Supplemental Table 4), indicate greater HbO values in controls compared to PWA.

In sum, during Semantic Matching, healthy controls showed canonical, left-lateralized hemodynamic responses across multiple language regions (IFGtri, IFGop, and MTG), whereas PWA demonstrated comparatively limited group-level activation. Between-group differences were most robust in left IFGop, where controls exhibited greater HbO than PWA during the later portion of the hemodynamic response.

#### 3.1.2. Rhyme Judgment

Changes in HbO and HbR for Rhyme Judgment across ROIs are depicted in Figure 4.

**Figure 4.**
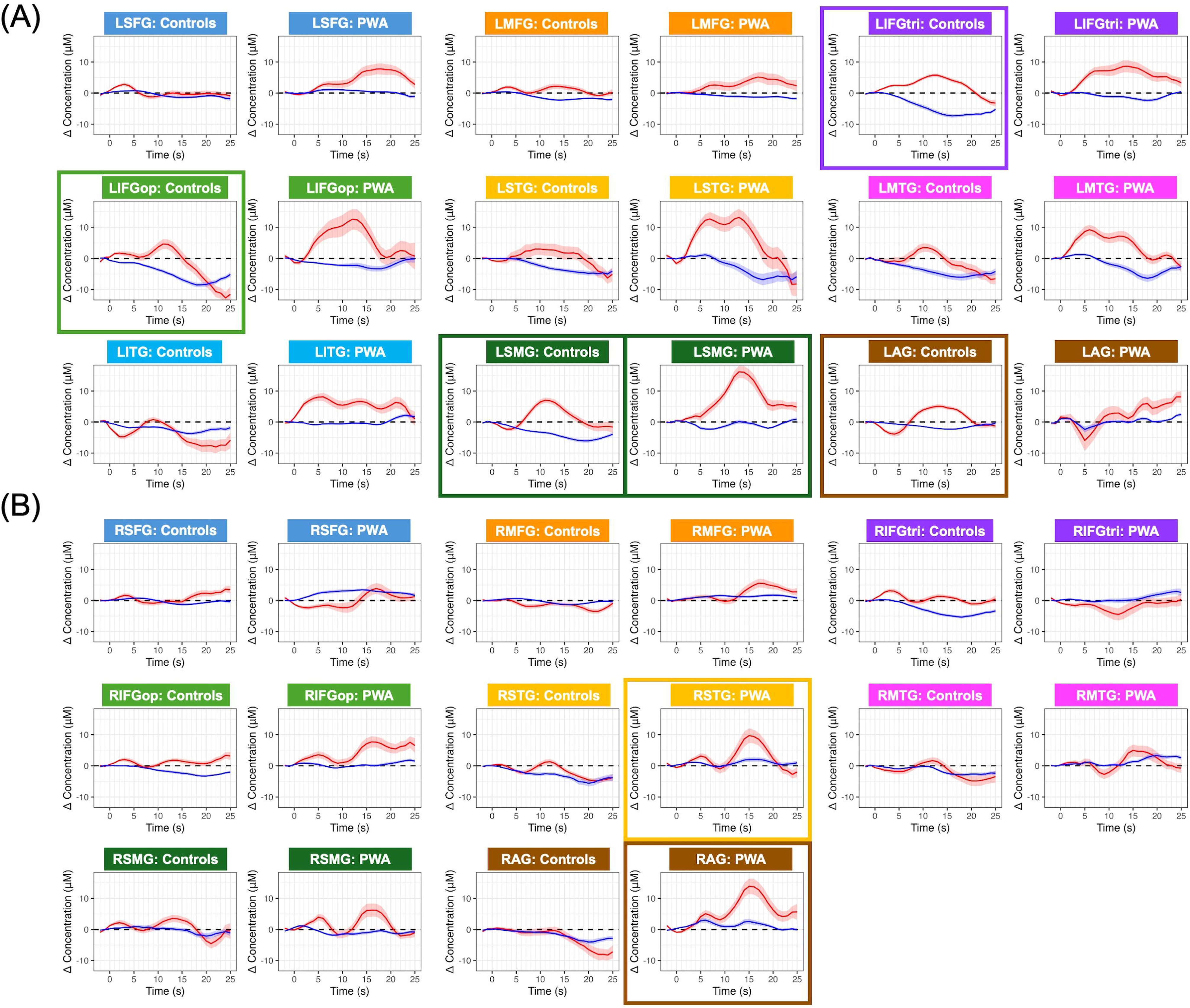
Changes in oxyhemoglobin (in red) and deoxyhemoglobin (in blue) for left hemisphere (A) and right hemisphere (B) regions of interest during Rhyme Judgment. For each region of interest (ROI), the left-side plot shows the group-averaged hemodynamic GLM output for controls, and the right-side plot shows the group-averaged hemodynamic GLM output for participants with aphasia. Solid color borders reflect ROIs for which a canonical and significant oxyhemoglobin time course was observed. Shaded ribbons represent within-group standard error around the mean time series. AG = angular gyrus, IFGtri/op = inferior frontal gyrus, pars triangularis/opercularis, ITG = inferior temporal gyrus, LH = left hemisphere, MFG = middle frontal gyrus, MTG = middle temporal gyrus, RH = right hemisphere, SFG = superior frontal gyrus, SMG = supramarginal gyrus, STG = superior temporal gyrus.

Table 4 reports significant omnibus model effects within each group. The full set of within-group Rhyme Judgment LMEM output for controls is reported in Supplemental Table 5 and in Supplemental Table 6 for PWA.

**Table 4.** Significant Within-Group Time Effects During Rhyme Judgment.

| Group | Hemoglobin | Region | *N* | *F* | *df* | *p* | *q* (FDR) |
| --- | --- | --- | --- | --- | --- | --- | --- |
| Controls | HbO |  |  |  |  |  |  |
|  |  | LIFGtri | 16 | 2.34 | 15, 225 | .004 | .020 |
|  |  | LIFGop | 16 | 2.3 | 15, 225 | .005 | .020 |
|  |  | LSMG | 16 | 3.61 | 15, 225 | < .001 | < .001 |
|  |  | LAG | 14 | 2.34 | 15, 195 | .004 | .020 |
|  | HbR |  |  |  |  |  |  |
|  |  | LMFG | 16 | 3.37 | 15, 225 | < .001 | < .001 |
|  |  | LIFGtri | 16 | 4.88 | 15, 225 | < .001 | < .001 |
|  |  | LIFGop | 16 | 3.36 | 15, 225 | < .001 | < .001 |
|  |  | LSMG | 16 | 2 | 15, 225 | .017 | .046 |
|  |  | RMFG | 16 | 2.35 | 15, 225 | .004 | .013 |
|  |  | RIFGtri | 16 | 2.36 | 15, 225 | .004 | .013 |
|  |  | RMTG | 16 | 1.96 | 15, 225 | .019 | .046 |
| PWA | HbO |  |  |  |  |  |  |
|  |  | LSMG | 11 | 3.01 | 15, 150 | < .001 | .005 |
|  |  | RSTG | 13 | 2.79 | 15, 180 | < .001 | .005 |
|  |  | RMTG | 13 | 1.73 | 15, 180 | .049 | .160 |
|  |  | RSMG | 13 | 1.93 | 15, 180 | .023 | .097 |
|  |  | RAG | 13 | 2.27 | 15, 180 | .006 | .033 |
|  | HbR |  |  |  |  |  |  |
|  |  | LSTG | 6 | 1.89 | 15, 75 | .038 | .216 |
|  |  | LMTG | 10 | 2.34 | 15, 135 | .005 | .089 |
|  |  | RSTG | 13 | 1.87 | 15, 180 | .029 | .216 |

Note. Only omnibus time effects with uncorrected $p < 0.05$ or FDR-corrected $q < 0.05$ are shown. Full model outputs are reported in Supplement Table 5 for controls and Supplemental Table 6 for participants with aphasia (PWA). AG = angular gyrus, HbO = oxyhemoglobin, HbR = deoxyhemoglobin, IFGtri/op = inferior frontal gyrus, pars triangularis/opercularis, ITG = inferior temporal gyrus, L = left, MFG = middle frontal gyrus, MTG = middle temporal gyrus, R = right, SFG = superior frontal gyrus, SMG = supramarginal gyrus, STG = superior temporal gyrus.

As a group, healthy controls demonstrated significant increases in HbO with corresponding significant decreases in HbR in left IFGtri from 8-18 s, left IFGop from 10-17 s, and left SMG from 10-13 s for both hemoglobin types (Table 4). Within left AG, the HbO response was canonically shaped and significant from 12-14 s, and while HbR retained the expected curve, the decrease was nonsignificant. Significant decreases in HbR were also observed in left MFG from 10-20 s, right MFG from 15-17 s, right IFGtri from 9-20 s, and right MTG from 15-20 s, but the HbO responses in each of these regions were nonsignificant with some regions exhibiting atypical HbO response morphology. As a group, PWA demonstrated significant increases in HbO without corresponding decreases in HbR in left SMG from 11-17 s, right STG from 14-17 s, and right AG from 13-18 s. There were no other significant findings for PWA after multiple comparison correction. The full set of Rhyme Judgment LMEM output for controls are reported in Supplemental Table 4 and within Supplemental Table 5 for PWA.

Significant findings from the between-group LMEMs are reported in Table 5, with full LMEM model output provided in Supplemental Table 7. Following correction for multiple comparisons, significant Group x Time interactions were observed for right STG and right MTG. Follow-up estimated marginal mean comparisons indicated greater HbO values in PWA than controls in right STG from 15-17 s at the uncorrected level. In right MTG, no individual time-point comparisons were significant either before or after correction.

**Table 5.** Significant Effects of Group and Group x Time During Rhyme Judgment.

| **Hemoglobin** | **ROI** | **Effect** | ***N* PWA** | ***N* Controls** | ***F*** | ***df*** | ***p*** | ***q*** |
| --- | --- | --- | --- | --- | --- | --- | --- | --- |
| HbO | RSTG | Group x Time | 13 | 16 | 2.70 | 15, 405 | < .001 | .010 |
| HbO | RMTG | Group x Time | 13 | 16 | 2.22 | 15, 405 | .006 | .047 |
| HbR | RIFGtri | Group x Time | 13 | 16 | 1.72 | 15, 405 | .045 | .375 |
| HbR | RMTG | Group x Time | 13 | 16 | 2.05 | 15, 405 | .012 | .196 |

Note. Only omnibus Group and Group x Time effects with uncorrected $p < 0.05$ or FDR-corrected $q < 0.05$ are shown. Full model outputs are reported in Supplement Table 7. HbO = oxyhemoglobin, HbR = deoxyhemoglobin, IFGtri = inferior frontal gyrus, pars triangularis, MTG = middle temporal gyrus, R = right, STG = superior temporal gyrus.

Overall, during Rhyme Judgment, healthy controls showed robust canonical hemodynamic responses in predominantly left-lateralized phonological regions (IFGtri, IFGop, SMG), whereas PWA demonstrated more limited activation with significant responses extending into right temporoparietal cortex. Between-group analyses identified differences in the temporal response profiles of right STG and MTG, although follow-up evidence for greater activation in PWA was limited and did not survive correction for multiple comparisons.

### 3.2. Behavioral Comparisons and Brain-Behavior Relationships Across PWA

#### 3.2.1. Comparisons between PWA and controls in ALM task performance

To contextualize the brain-behavior findings within PWA, we first evaluated how ALM behavioral performance varied between groups for the two task conditions (i.e., experimental semantic/rhyme trials versus control perceptual judgment trials). As shown in the top panel of Figure 5 and reported in Table 6, accuracy differed significantly by both group and condition for Semantic Matching. However, the group by condition interaction was not significant, indicating that the effect of condition on accuracy did not differ noticeably between controls and PWA. A similar pattern emerged for RT, with significant main effects of group and condition, but the group by condition interaction only approached significance. Mean trial difficulty also differed significantly between groups and conditions without a significant interaction. Thus, across Semantic Matching metrics, there was no evidence that condition affected PWA and controls differently. Rather, PWA had lower accuracy, slower response times, and lower mean trial difficulty for both conditions.

**Figure 5.**
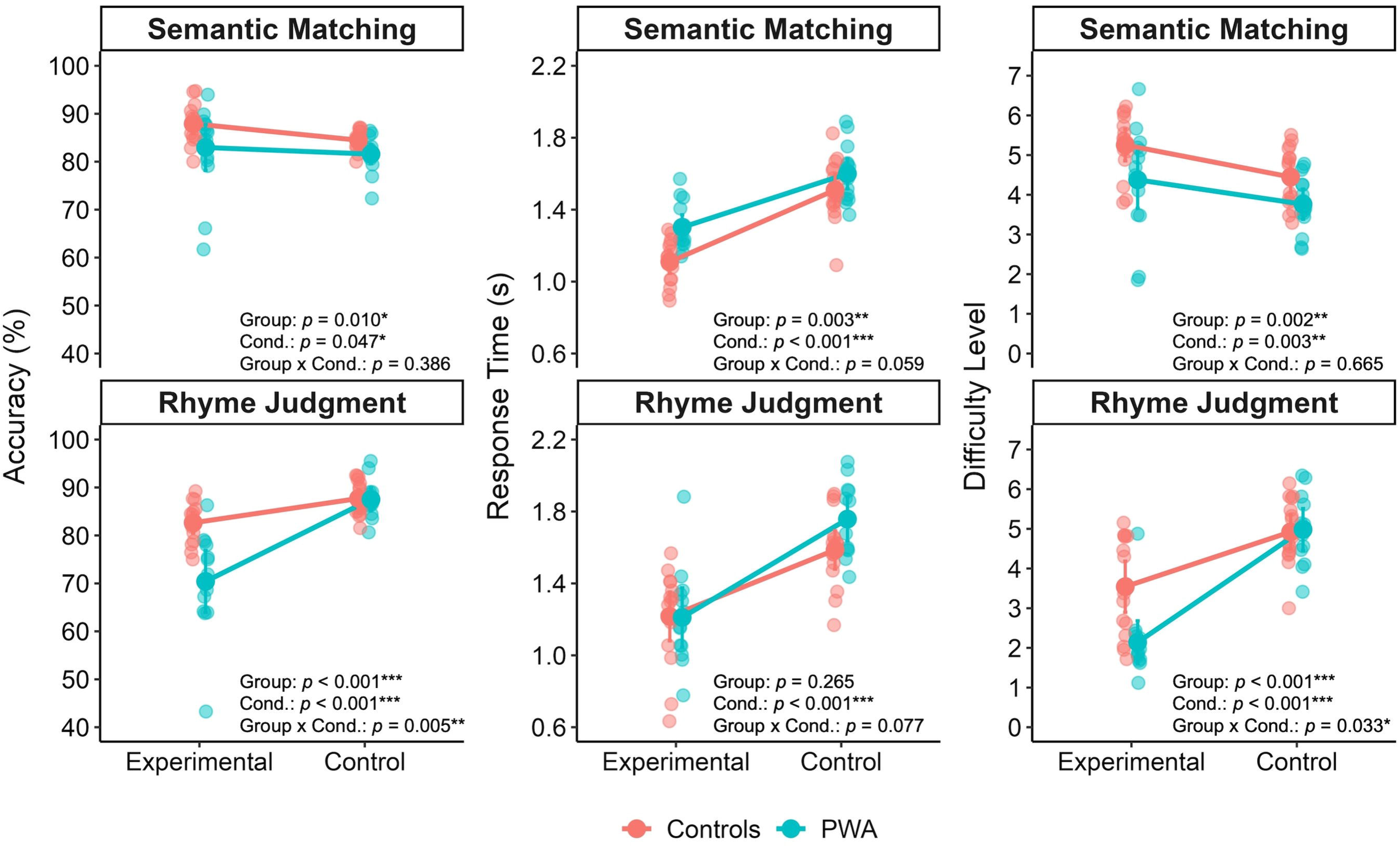
Behavioral results from both fNIRS tasks. In each plot, the experimental semantic/rhyme decision condition is plotted to the left and the perceptual judgment condition is plotted to the right. Data reflect the mean accuracy (left column), mean reaction time (RT, middle column), and mean difficulty level (right column) of trials across each experiment. Cond. = condition, PWA = participants with aphasia.

**Table 6.** Between-Group and Between-Condition Comparisons of Task Performance.

| Task | Variable | Effect | *F* | *df* | *p* | $\eta^2G$ |
| --- | --- | --- | --- | --- | --- | --- |
| Semantic Matching | Accuracy | Group | 7.50 | 1, 29 | .010 | 0.135 |
|  |  | Condition | 4.29 | 1, 29 | .047 | 0.055 |
|  |  | Group x Condition | 0.77 | 1, 29 | .386 | 0.010 |
|  | RT | Group | 10.12 | 1, 29 | .003 | 0.207 |
|  |  | Condition | 182.85 | 1, 29 | < .001 | 0.613 |
|  |  | Group x Condition | 3.86 | 1, 29 | .059 | 0.032 |
|  | Mean trial difficulty | Group | 11.22 | 1, 29 | .002 | 0.169 |
|  |  | Condition | 10.24 | 1, 29 | .003 | 0.144 |
|  |  | Group x Condition | 0.19 | 1, 29 | .665 | 0.003 |
| Rhyme Judgment | Accuracy | Group | 27.94 | 1, 29 | < .001 | 0.216 |
|  |  | Condition | 32.45 | 1, 29 | < .001 | 0.469 |
|  |  | Group x Condition | 9.55 | 1, 29 | .005 | 0.206 |
|  | RT | Group | 1.29 | 1, 29 | .265 | 0.032 |
|  |  | Condition | 90.90 | 1, 29 | < .001 | 0.504 |
|  |  | Group x Condition | 3.39 | 1, 29 | .077 | 0.037 |
|  | Mean trial difficulty | Group | 18.05 | 1, 29 | < .001 | 0.113 |
|  |  | Condition | 42.13 | 1, 29 | < .001 | 0.558 |
|  |  | Group x Condition | 5.05 | 1, 29 | .033 | 0.131 |

Note. Follow-up between-group comparisons for significant interaction effects are reported in the manuscript text. $\eta^2G$ = generalized eta squared, RT = reaction time.

As shown in the bottom panel of Figure 5 and reported in the lower portion of Table 6, a different pattern emerged for the Rhyme Judgment task. Accuracy varied by group and condition, and the interaction term was significant. Follow-up comparisons indicated that controls and PWA did not differ in accuracy for the control condition (at 87.7% vs. 87.5%, *t*_27_ = 0.13, *q* = 0.897), but controls were significantly more accurate than PWA for the experimental condition (at 82.6% vs. 70.4%, *t*_27_ = 4.22, *q* < 0.001). In contrast, RT differed greatly across conditions, but there was no group difference nor significant group by condition interaction. Like accuracy, mean trial difficulty showed significant effects of group and condition, and the interaction term was significant. With mean experimental condition difficulty at 3.54 for controls and 2.14 for PWA, the between-group difference was evident for rhyme judgments (*t*_27_ = 3.47, *q* = 0.002), whereas controls and PWA had similar mean difficulty (4.92 vs. 4.98) for the perceptual decision condition (*t*_27_ =-0.19, *q* = 0.850).

#### 3.2.2. Relationships between averaged HbO and ALM task performance

As shown in Table 7, before FDR correction, lower standardized Semantic Matching scores were significantly associated with greater damage to left ventral stream regions but not with damage to left dorsal stream structures. Higher Semantic Matching scores—indicative of more control-like behavioral performance—were associated with greater activation of left dorsal stream regions but not with left ventral, right ventral or right dorsal regions.

**Table 7.**
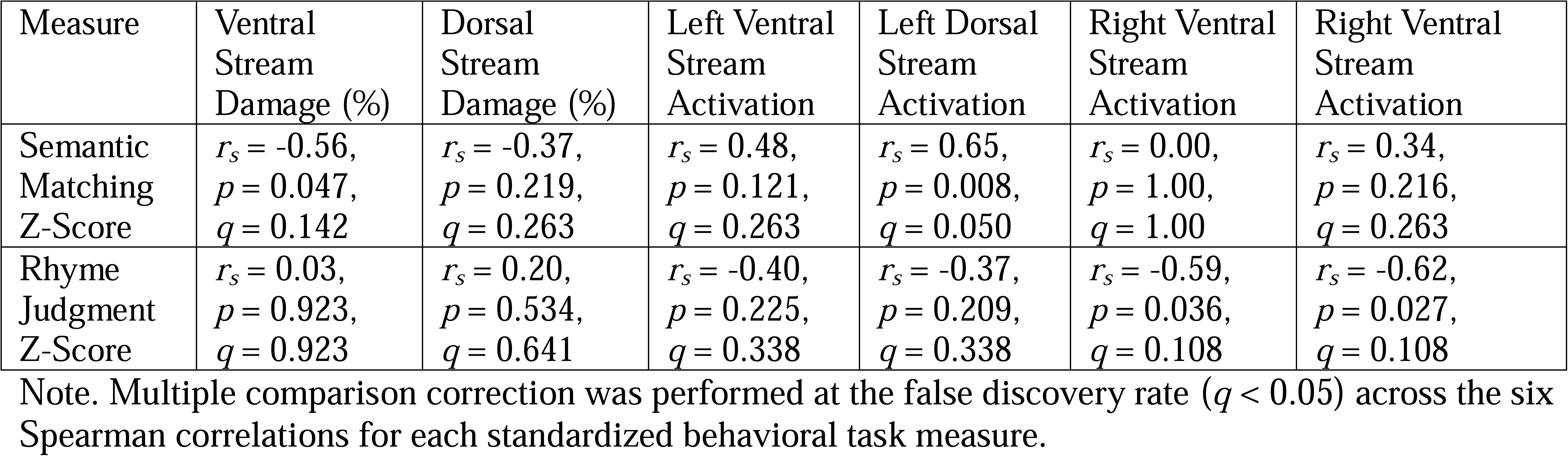
Correlations Between Standardized Task Performance and Dual-Stream Measures Within Participants With Aphasia.

Rhyme Judgment performance in PWA was not significantly associated with damage to either left ventral or dorsal stream regions. Likewise, performance was not significantly related to the degree of activity within the left ventral or dorsal streams. In contrast, more control-like behavioral performance on Rhyme Judgment was significantly associated with lower activation of right ventral and right dorsal cortex, although neither finding survived correction for multiple comparisons (Table 7).

## 4. Discussion

In this study, we utilized fNIRS to examine the extent to which adaptive semantic and phonological language-mapping tasks recruit regions of the dual-stream language network in adults with and without aphasia, and whether task-specific performance in PWA relates to structural damage and functional recruitment within these networks. In neurologically healthy adults, the two tasks produced largely distinct activation patterns consistent with predictions from dual-stream models, providing initial support for the use of fNIRS with the ALM paradigms. In contrast, PWA demonstrated substantial inter-individual variability and relatively few significant regions at the group level, with less apparent segregation of dorsal and ventral stream activity. Brain-behavior associations further suggested that individual differences in both lesion distribution and recruitment of surviving cortical tissue contributed to language performance, although these findings should be interpreted cautiously given the small sample.

### 4.1. fNIRS-Based Language Mapping in Neurologically Healthy Adults Aligns with Dual-Stream Organization

A critical precursor to using functional imaging to study post-stroke language reorganization is demonstrating that the experimental paradigms and imaging modality elicit expected patterns of functional organization in neurologically healthy adults (Wilson & Schneck, 2021). The present findings indicate that within probed regions, the ALM tasks produce broadly comparable patterns to fMRI when implemented with fNIRS. Semantic Matching preferentially recruited ventral-stream regions, with prominent activity in the left superior and middle temporal cortex, whereas Rhyme Judgment produced stronger recruitment of dorsal regions, particularly the left inferior frontal and inferior parietal cortex. Semantic Matching elicited widespread left temporal activity, with the strongest and most consistent response observed in left MTG and weaker recruitment in neighboring STG and posterior ITG. Given the spatial resolution of fNIRS, this pattern may reflect a response centered in the middle temporal cortex, consistent with meta-analytic and review evidence implicating mid to posterior MTG in semantic representation and controlled retrieval, respectively (Binder et al., 2009; Hodgson et al., 2021; Jackson, 2021; Lambon Ralph et al., 2017). Likewise, recruitment of left SMG during Rhyme Judgment accords with functional neuroimaging evidence linking this region to phonological encoding (e.g., Devlin et al., 2003; Gitelman et al., 2005; McDermott et al., 2003; Oberhuber et al., 2016; Price et al., 1997) as well as lesion-symptom mapping studies associating damage to this region with impaired phonological recognition in PWA (Alyahya et al., 2020a; Butler et al., 2014; Dell et al., 2013; Fridriksson et al., 2018; Pillay et al., 2014; Schumacher et al., 2019; Schwartz et al., 2012). Although the ROI-level analysis also implicated left AG during Rhyme Judgment, inspection of the underlying channel-level responses suggested that this effect was centered primarily over left SMG with some extension into the anterior portion of AG.

The principal area of overlap between tasks was the left inferior cortex, as left IFGtri and IFGop were the only ROIs significantly recruited during both Semantic Matching and Rhyme Judgment in healthy controls. Channel-level responses during Rhyme Judgment appeared concentrated in more posterior portions of left IFG, consistent with evidence for an anteroventral-to-posterodorsal gradient, whereby relatively anterior/ventral areas are preferentially engaged during semantic processing and posterior/dorsal areas during phonological processing (Devlin et al., 2003; Gough et al., 2005; Hartwigsen et al., 2020; Klaus & Hartwigsen, 2019; McDermott et al., 2003; Poldrack et al., 1999). Semantic Matching, however, recruited both anterior and posterior IFG, consistent with prior findings of the ALM paradigms from fMRI (Wilson et al., 2018; Yen et al., 2019). Importantly, overlap at the relatively coarse ROI level does not necessarily imply an absence of functional specialization. While functionally distinct linguistic and domain-general populations are spatially interdigitated within the frontal cortex (Fedorenko et al., 2011, 2012, 2013; Fedorenko & Blank, 2020) the spatial resolution of our sparse fNIRS array is insufficient to distinguish neighboring populations within a single anatomical ROI. Thus, spatially overlapping activations could reflect common or different linguistic demands across tasks, domain-general demands such as maintaining task rules or selecting among competing responses, or a combination of these processes (Fedorenko et al., 2012). Incorporating a separate adaptive nonlinguistic task into future investigations could help distinguish domain-specific from domain-general contributions to fNIRS activity within left IFG.

Finally, activation pattern differences between our healthy control sample and those reported by Wilson and colleagues (Wilson et al., 2018; Yen et al., 2019) primarily lie in subcortical regions (e.g., left hippocampus) and extracerebral brain structures (e.g., right cerebellum) that could not be reached by the fNIRS probes. As a key exception, the channels overlying left SFG that showed a weak response in our Semantic Matching data are spatially approximated (although anterior and inferior) to peaks in left SFG that showed a significant response in Wilson et al. (2018) and Yen et al. (2019). Interestingly, in Wilson et al. (2018), the magnitude and extent of activation within this region increased with a more difficult compared to easier version of the task, which the authors posited could reflect the heightened task challenge. Although purely speculative, the lack of significant left SFG recruitment for either task in our control sample may indicate task difficulty was appropriately titrated to individual subjects.

4.2. PWA Show Heterogeneous and Less Segregated Patterns of Dual-Stream Recruitment In contrast to the relatively clear task differentiation observed in controls, activation patterns in PWA were considerably more heterogeneous. Perhaps the most striking group-level finding was the limited number of regions demonstrating statistically reliable activity despite the presence of plausible hemodynamic responses in several ROIs. For example, no region demonstrated a significant HbO increase during Semantic Matching at the group level, despite prior work showing robust single-subject activation in PWA with the fMRI version of this paradigm (Wilson et al., 2018). This discrepancy likely reflects, at least in part, the considerable heterogeneity in lesion location and hemodynamic responses characteristic of post-stroke aphasia (Bonakdarpour et al., 2007), compounded by the limited statistical power of the present sample.

Nevertheless, several features of the average responses were consistent with preservation of canonical language-network organization. During Semantic Matching, left IFGtri and left MTG—regions frequently implicated in semantic retrieval and control (Lambon Ralph et al., 2017)—showed relatively canonical, albeit nonsignificant, hemodynamic responses. Similarly, Rhyme Judgment elicited its strongest response in left SMG, consistent with both our findings in neurologically healthy adults and the established role of the inferior parietal cortex in phonological processing. Left SMG showed little evidence of recruitment during Semantic Matching, providing some indication that phonological specialization within this region may be preserved following stroke. Canonically shaped responses were also evident in both left IFGtri and IFGop during Rhyme Judgment, although substantial between-participant variability prevented these effects from reaching significance.

At the same time, the data from PWA did not reveal a clear group-level segregation of dorsal and ventral stream activity. As a key example, Rhyme Judgment elicited responses not only within expected dorsal regions but also smaller group-level effects throughout the left temporal cortex, including left STG, MTG, and ITG. Both tasks also elicited canonical (albeit nonsignificant) responses within left SFG. Thus, unlike controls, it is possible that the difficulty of the tasks was not entirely controlled across PWA, particularly for Rhyme Judgment. Indeed, an important consideration for Rhyme Judgment is that the adaptive nature of the task may have resulted in qualitatively different processing demands across participants. On average, PWA progressed to substantially lower difficulty levels than controls, and most completed a high proportion of trials at the two lowest levels. At these levels, orthographic similarity between nonword pairs can sometimes provide sufficient information to make a response without fully converting orthographic representations to phonology and segmenting the resulting phonological forms. Higher-level trials, in contrast, more consistently require phonological analysis because orthographic similarity is less informative. Consequently, the broader activation pattern observed in PWA may partly reflect greater reliance from some participants on visual or orthographic strategies and/or attempts to use semantics to process nonword stimuli, rather than phonological processing per se. This issue also complicates direct interpretation of between-group differences in activation despite the benefits of adaptive difficulty for equating overall task performance.

Future work using the auditory version of Rhyme Judgment could help ensure that successful performance requires phonological encoding and comparison across difficulty levels.

Another salient finding from the group-level activation results in PWA was the relatively limited significant engagement of right hemisphere regions during either task. Meta-analyses of prior fMRI studies indicate that, across language tasks, PWA engage right hemisphere cortex— particularly right frontal regions such as IFG and the insula—more strongly than neurologically healthy adults (LaCroix et al., 2021; Stefaniak et al., 2021; Turkeltaub et al., 2011). In the present study, Rhyme Judgment elicited small increases in HbO across several right hemisphere regions, but only right STG and AG showed significant changes, neither of which was accompanied by a corresponding decrease in HbR. Semantic Matching showed a markedly different pattern: like controls, PWA exhibited trends towards decreased HbO with co-occurring increased HbR across several right ROIs, consistent with deactivation. This pattern was somewhat unexpected given that the ventral language pathway is generally considered less strongly left-lateralized than the dorsal pathway (Hickok & Poeppel, 2007). Importantly, task differences remained largely unchanged when Semantic Matching analyses were restricted to participants who completed both tasks (Supplemental Figure 1), suggesting that differences in cohort composition were unlikely to account for the contrasting right hemisphere responses. At the same time, the limited right hemisphere recruitment observed during Semantic Matching coheres with recent findings from Turkeltaub et al. (2026), who used the same adaptive Semantic Matching paradigm in fMRI and found that only 14 of 76 PWA exhibited greater right hemisphere activity than controls, and at a group level, right hemisphere recruitment was modest compared to left hemisphere engagement.

### 4.3. Brain-Behavior Relationships Suggest Contributions of Both Structural Integrity and Functional Recruitment

Individual differences in lesion distribution and functional recruitment provided additional insight into the heterogeneous group-level findings in PWA. Semantic Matching performance was poorer in individuals with greater damage to the left ventral stream, consistent with the central role of ventral temporal regions in semantic processing. At the same time, more control-like semantic performance was associated with greater activation of left dorsal stream regions. Together, these findings raise the possibility that successful semantic performance after stroke depends both on preservation of canonical ventral circuitry and on recruitment of additional surviving portions of the left hemisphere language network, findings that appear to align with those reported by Pillay et al. (2022) and Stefaniak and colleagues’ (2020) variable neuro-displacement theory. This theory posits that quiescent brain regions may be a part of the healthy network and are only upregulated in instances when the system is under stress, as would be the case after stroke.

The relationship between dorsal recruitment and semantic performance should not, however, be interpreted straightforwardly as compensatory takeover of semantic processing by the dorsal stream. A variable neuro-displacement account across language subnetworks (Stefaniak et al., 2020) would predict that increasing damage to canonical ventral regions should be accompanied by greater recruitment of alternative dorsal regions. Instead, a follow-up analysis showed that greater left ventral damage was associated with *lower* left dorsal activation (*r_s_* =-0.42, *p* = 0.152). Therefore, one possibility is that recruitment of dorsal regions during Semantic Matching depends on broader preservation of the left language network rather than being triggered specifically by destruction of ventral semantic tissue. Under this interpretation, PWA with relatively preserved left hemisphere network resources may be better able to engage both canonical and ancillary language regions, whereas individuals with more extensive damage have less surviving tissue available to support either process.

For Rhyme Judgment, behavioral performance was generally negatively associated with bilateral dual-stream recruitment, with the strongest relationships indicating greater right ventral and dorsal activity in individuals with poorer phonological performance. The role of the right hemisphere in aphasia recovery has long been debated, with increased contralesional activity interpreted alternatively as beneficial compensation (e.g., Saur et al., 2006; Winhuisen et al., 2005), inefficient or unsuccessful recruitment (e.g., Postman-Caucheteux et al., 2010), or a consequence of the extent and distribution of left hemisphere damage (e.g., Griffis et al., 2017; Skipper-Kallal et al., 2017). The present findings suggest individuals who struggled most with the Rhyme Task engaged usually quiescent right hemisphere regions, perhaps due to greater processing demands or in an attempt to recruit additional neural resources when canonical left hemisphere systems were insufficient. However, given the small sample and correlational approach, distinguishing potential causal detriment from true functional compensation from the effects of residual network integrity will require replication in larger cohorts and methods capable of explicitly modeling the relationship among lesion burden, activation, and behavior.

### 4.4. Study Limitations and Future Directions

The preliminary nature of these findings should be emphasized. Foremost, the PWA sample was small, limiting statistical power for both group-level activation and brain-behavior analyses. Lesions were also not evenly distributed across canonical language regions, with damage concentrated particularly in the inferior parietal cortex (Figure 2), and aphasia severity was skewed toward the milder end of the spectrum (Table 1). Indeed, some participants scored outside the aphasic range on the WAB-R Aphasia Quotient despite continuing to report meaningful language challenges after stroke. Replication in a larger sample encompassing a broader range of aphasia severity and lesion distributions—including greater representation of frontal lesions—will be necessary to determine the generalizability of the observed patterns.

Additional limitations relate to fNIRS itself and to the array used in the present study.

The limited penetration depth of fNIRS precludes direct measurement of subcortical and deeper cortical structures implicated in language processing and recovery, while the relatively sparse optode array provided limited coverage of extrasylvian cortex. Higher-density arrays could improve spatial specificity, extend cortical coverage, and permit inclusion of control regions not expected to activate during language tasks. High-density diffuse optical tomography may additionally improve localization, although depth limitations relative to fMRI would remain.

Finally, a central motivation of this study was to evaluate the feasibility of fNIRS as an accessible alternative to fMRI for language mapping in adults with and without aphasia. An important next step is therefore to establish the reliability of task-evoked responses at the individual level. This could include split-half reliability within a run (as performed by Wilson et al., 2018), reliability across repeated runs within a session, and test-retest reliability across longer intervals. Demonstrating robust within-person reliability is particularly important for aphasia research, in which functional imaging is frequently used to characterize treatment-related neuroplasticity or longitudinal recovery. If adequate reliability can be established, fNIRS-based adaptive language mapping may provide a useful platform not only for tracking regional activation over time but also for investigating individual functional-network organization and cortical connectome-based signatures of language recovery.

## 5. Conclusions

Altogether, we found the expected dual-route architecture when adaptive language mapping was performed using fNIRS in neurologically healthy adults. In contrast, the group-level and individual-differences findings in PWA suggest that the functional organization of semantic and phonological processing after stroke cannot be characterized simply as preservation versus loss of canonical dual-stream architecture. Some regions—particularly the left temporal cortex for semantic processing and the left inferior parietal cortex for phonological processing— showed evidence of retaining expected functional roles. At the same time, activation was more spatially distributed and variable across PWA, and behavioral performance was related to recruitment outside the regions most strongly associated with each process in healthy adults.

These findings are consistent with a model in which post-stroke language performance reflects the combined influence of residual specialization, lesion-dependent constraints on available network resources, and flexible recruitment of surviving tissue. Overall, the results indicate fNIRS may be a viable alternative to fMRI for language mapping in older adults with and without aphasia, but replication with a more extensive fNIRS array and larger samples is warranted.

## Author Contributions: CRediT

**Erin Meier:** Conceptualization, Funding acquisition, Data curation, Formal analysis, Investigation, Methodology, Project administration, Resources, Supervision, Visualization, Writing – original draft; **Veronica Fletcher:** Data curation, Formal analysis, Investigation, Methodology, Writing – Review & editing; **Caela Hung:** Formal analysis, Validation, Writing – Review & editing; **Esprit Ange Andraos:** Data curation, Investigation, Writing – Review & editing; **Leanna Ugent:** Data curation, Investigation, Writing – Review & editing; **Gengchen Wei:** Data curation, Investigation, Writing – Review & editing; **David Lin:** Resources, Writing – Review & editing; **Meryem Yücel:** Validation, Writing – Review & editing.

## Data Availability

All data produced from the present study are available on Open Science Framework (Meier, 2026).

https://osf.io/zk4j5/overview

## Acknowledgments

We are grateful to other members of The Aphasia Network Laboratory at Northeastern University who supported participant recruitment, data collection, and data analysis efforts. We are also appreciative of our participants for their time and effort in completing study procedures.

