## Supplementary Material for "Functional near infrared spectroscopy-based dual-stream adaptive language mapping in adults with and without post-stroke aphasia"

Supplemental Table 1. Region assignments and retention status of long-distance fNIRS channels

| ROI | Channel assignment (MNI coordinates) | Assigned Pathway | Semantic Matching | | | | | Rhyme Judgment | | | | |
| --- | --- | --- | --- | --- | --- | --- | --- | --- | --- | --- | --- | --- |
|  |  |  | Controls | | PWA | | | Controls | | PWA | | |
|  |  |  | Retained | Pruned | Retained | Pruned | Lesioned | Retained | Pruned | Retained | Pruned | Lesioned |
| LSFG | Ch1 (-10 55 22); Ch2 (-22 46 21) | N/A | 32 | 0 | 26 | 0 | 0 | 32 | 0 | 24 | 0 | 0 |
| LMFG | Ch3 (-30 59 6);  Ch4 (-38 37 20) | Dorsal | 32 | 0 | 24 | 0 | 2 | 31 | 1 | 23 | 0 | 1 |
| LIFGtri | Ch5 (-33 22 24); Ch6 (-40 16 25); Ch7 (-40 49 6);  Ch8 (-35 22 22);  Ch9 (-39 47 3); Ch10 (-45 28 19); Ch11 (-55 13 13) | Dorsal/Ventral | 112 | 0 | 67 | 0 | 24 | 112 | 0 | 67 | 0 | 17 |
| LIFGop | Ch12 (-48 7 25) | Dorsal/Ventral | 16 | 0 | 9 | 0 | 4 | 16 | 0 | 8 | 0 | 4 |
| LSTG | Ch13 (-53 -14 9) | Ventral | 16 | 0 | 9 | 0 | 4 | 16 | 0 | 9 | 0 | 3 |
| LMTG | Ch14 ( -62 -1 -6); Ch15 (-43 -38 8); Ch16 ( -49 -26 -6) | Ventral | 48 | 0 | 20 | 0 | 19 | 47 | 1 | 20 | 0 | 16 |
| LITG | Ch17 ( -47 -46 -6) | Ventral | 16 | 0 | 5 | 0 | 8 | 16 | 0 | 5 | 0 | 7 |
| LSMG | Ch18 (-56 -24 35); Ch19 (-46 -24 25); Ch20 (-56 -52 26) | Dorsal | 45 | 3 | 24 | 0 | 15 | 45 | 3 | 24 | 0 | 12 |
| LAG | Ch21 (-53 -53 32) | Dorsal | 16 | 0 | 6 | 0 | 7 | 16 | 0 | 6 | 0 | 6 |
| RSFG | Ch23 (16 69 28) | N/A | 16 | 0 | 15 | 0 | 0 | 16 | 0 | 13 | 0 | 0 |
| RMFG | Ch24 (33 57 28); Ch25 (40 67 9); Ch26 (42 32 17) | Dorsal | 48 | 0 | 45 | 0 | 0 | 48 | 0 | 39 | 0 | 0 |
| RIFGtri | Ch27 (44 46 2); Ch28 (46 24 19); Ch29 (55 53 1); Ch30 (56 27 16) | Dorsal/Ventral | 63 | 1 | 60 | 0 | 0 | 61 | 3 | 52 | 0 | 0 |
| RIFGop | Ch31 (45 26 32); Ch32 (55 17 31) | Dorsal/Ventral | 31 | 1 | 30 | 0 | 0 | 31 | 1 | 26 | 0 | 0 |
| RSTG | Ch36 (63 -26 23); Ch37 (65 -11 9); Ch38 (72 -43 12) | Ventral | 47 | 1 | 45 | 0 | 0 | 44 | 4 | 39 | 0 | 0 |
| RMTG | Ch39 (53 -26 -8); Ch40 (58 -46 -5); Ch41 (43 -56 12) | Ventral | 46 | 2 | 45 | 0 | 0 | 46 | 2 | 39 | 0 | 0 |
| RSMG | Ch42 (50 -23 32) | Dorsal | 15 | 1 | 15 | 0 | 0 | 15 | 1 | 13 | 0 | 0 |
| RAG | Ch43 (45 -46 30); Ch44 (59 -50 25) | Dorsal | 31 | 1 | 30 | 0 | 0 | 31 | 1 | 26 | 0 | 0 |
| Total Channels | |  | 630 | 10 | 475 | 0 | 83 | 623 | 17 | 433 | 0 | 66 |

Notes. Values in the retained, pruned, and lesioned columns reflect channel counts for each group. AG = angular gyrus, Ch = channel, IFGtri/op = inferior frontal gyrus, pars triangularis/opercularis, ITG = inferior temporal gyrus, L = left, MFG = middle frontal gyrus, MTG = middle temporal gyrus, PWA = participants with aphasia, R = right, ROI = region of interest, SFG = superior frontal gyrus, SMG = supramarginal gyrus, STG = superior temporal gyrus.

Supplemental Table 2. Linear mixed effects models results for Semantic Matching for healthy controls

| **Region** | ***N*** | ***F*** | ***df*** | **Effect / time** | ***t*** | **EMM (µM)** | **SE (µM)** | ***p*** | ***q* (FDR)** |
| --- | --- | --- | --- | --- | --- | --- | --- | --- | --- |
| **HbO** |  |  |  |  |  |  |  |  |  |
| **LSFG** | **16** | **0.87** | **15, 225** | **Overall time effect** |  |  |  | **.604** | **.765** |
| **LMFG** | **16** | **0.43** | **15, 225** | **Overall time effect** |  |  |  | **.969** | **.999** |
| **LIFGtri** | **16** | **3.18** | **15, 225** | **Overall time effect** |  |  |  | **< .001** | **.002** |
|  |  |  | 15 | 5 s | 0.96 | 3.28 | 3.43 | .177 | .211 |
|  |  |  | 15 | 6 s | 1.51 | 5.17 | 3.43 | .076 | .111 |
|  |  |  | 15 | 7 s | 1.92 | 6.57 | 3.43 | .037 | .111 |
|  |  |  | 15 | 8 s | 2.04 | 7.00 | 3.43 | .030 | .111 |
|  |  |  | 15 | 9 s | 2.01 | 6.90 | 3.43 | .031 | .111 |
|  |  |  | 15 | 10 s | 1.87 | 6.40 | 3.43 | .041 | .111 |
|  |  |  | 15 | 11 s | 1.72 | 5.91 | 3.43 | .053 | .111 |
|  |  |  | 15 | 12 s | 1.64 | 5.62 | 3.43 | .061 | .111 |
|  |  |  | 15 | 13 s | 1.61 | 5.53 | 3.43 | .064 | .111 |
|  |  |  | 15 | 14 s | 1.65 | 5.64 | 3.43 | .060 | .111 |
|  |  |  | 15 | 15 s | 1.66 | 5.70 | 3.43 | .059 | .111 |
|  |  |  | 15 | 16 s | 1.53 | 5.26 | 3.43 | .073 | .111 |
|  |  |  | 15 | 17 s | 1.29 | 4.43 | 3.43 | .108 | .144 |
|  |  |  | 15 | 18 s | 0.93 | 3.18 | 3.43 | .185 | .211 |
|  |  |  | 15 | 19 s | 0.48 | 1.64 | 3.43 | .319 | .341 |
|  |  |  | 15 | 20 s | 0.03 | 0.11 | 3.43 | .487 | .487 |
| **LIFGop** | **16** | **2.52** | **15, 225** | **Overall time effect** |  |  |  | **.002** | **.010** |
|  |  |  | 15 | 5 s | 0.62 | 3.25 | 5.21 | .271 | .280 |
|  |  |  | 15 | 6 s | 1.05 | 5.46 | 5.21 | .156 | .178 |
|  |  |  | 15 | 7 s | 1.36 | 7.09 | 5.21 | .097 | .129 |
|  |  |  | 15 | 8 s | 1.51 | 7.85 | 5.21 | .076 | .121 |
|  |  |  | 15 | 9 s | 1.53 | 7.96 | 5.21 | .074 | .121 |
|  |  |  | 15 | 10 s | 1.45 | 7.58 | 5.21 | .083 | .121 |
|  |  |  | 15 | 11 s | 1.46 | 7.61 | 5.21 | .083 | .121 |
|  |  |  | 15 | 12 s | 1.65 | 8.60 | 5.21 | .060 | .121 |
|  |  |  | 15 | 13 s | 1.90 | 9.93 | 5.21 | .038 | .121 |
|  |  |  | 15 | 14 s | 2.14 | 11.17 | 5.21 | .025 | .116 |
|  |  |  | 15 | 15 s | 2.30 | 11.97 | 5.21 | .018 | .116 |
|  |  |  | 15 | 16 s | 2.27 | 11.85 | 5.21 | .019 | .116 |
|  |  |  | 15 | 17 s | 2.05 | 10.71 | 5.21 | .029 | .116 |
|  |  |  | 15 | 18 s | 1.65 | 8.59 | 5.21 | .060 | .121 |
|  |  |  | 15 | 19 s | 1.14 | 5.92 | 5.21 | .137 | .169 |
|  |  |  | 15 | 20 s | 0.60 | 3.11 | 5.21 | .280 | .280 |
| **LSTG** | **16** | **2.13** | **15, 225** | **Overall time effect** |  |  |  | **.010** | **.033** |
|  |  |  | 15 | 5 s | 1.47 | 9.21 | 6.28 | .082 | .109 |
|  |  |  | 15 | 6 s | 1.89 | 11.86 | 6.28 | .039 | .070 |
|  |  |  | 15 | 7 s | 2.20 | 13.81 | 6.28 | .022 | .067 |
|  |  |  | 15 | 8 s | 2.40 | 15.05 | 6.28 | .015 | .067 |
|  |  |  | 15 | 9 s | 2.45 | 15.36 | 6.28 | .014 | .067 |
|  |  |  | 15 | 10 s | 2.34 | 14.68 | 6.28 | .017 | .067 |
|  |  |  | 15 | 11 s | 2.18 | 13.70 | 6.28 | .023 | .067 |
|  |  |  | 15 | 12 s | 2.11 | 13.25 | 6.28 | .026 | .067 |
|  |  |  | 15 | 13 s | 2.05 | 12.87 | 6.28 | .029 | .067 |
|  |  |  | 15 | 14 s | 1.93 | 12.09 | 6.28 | .037 | .070 |
|  |  |  | 15 | 15 s | 1.75 | 10.99 | 6.28 | .050 | .080 |
|  |  |  | 15 | 16 s | 1.50 | 9.44 | 6.28 | .077 | .109 |
|  |  |  | 15 | 17 s | 1.24 | 7.75 | 6.28 | .118 | .145 |
|  |  |  | 15 | 18 s | 0.96 | 6.05 | 6.28 | .175 | .200 |
|  |  |  | 15 | 19 s | 0.67 | 4.19 | 6.28 | .257 | .275 |
|  |  |  | 15 | 20 s | 0.33 | 2.08 | 6.28 | .372 | .372 |
| **LMTG** | **16** | **2.89** | **15, 225** | **Overall time effect** |  |  |  | **< .001** | **.003** |
|  |  |  | 15 | 5 s | 2.31 | 12.08 | 5.23 | .018 | .029 |
|  |  |  | 15 | 6 s | 2.78 | 14.57 | 5.23 | .007 | .018 |
|  |  |  | 15 | 7 s | 3.03 | 15.87 | 5.23 | .004 | .018 |
|  |  |  | 15 | 8 s | 3.10 | 16.20 | 5.23 | .004 | .018 |
|  |  |  | 15 | 9 s | 3.11 | 16.25 | 5.23 | .004 | .018 |
|  |  |  | 15 | 10 s | 3.00 | 15.69 | 5.23 | .004 | .018 |
|  |  |  | 15 | 11 s | 2.84 | 14.89 | 5.23 | .006 | .018 |
|  |  |  | 15 | 12 s | 2.72 | 14.25 | 5.23 | .008 | .018 |
|  |  |  | 15 | 13 s | 2.56 | 13.41 | 5.23 | .011 | .022 |
|  |  |  | 15 | 14 s | 2.31 | 12.09 | 5.23 | .018 | .029 |
|  |  |  | 15 | 15 s | 1.95 | 10.22 | 5.23 | .035 | .051 |
|  |  |  | 15 | 16 s | 1.48 | 7.73 | 5.23 | .080 | .107 |
|  |  |  | 15 | 17 s | 1.03 | 5.38 | 5.23 | .160 | .197 |
|  |  |  | 15 | 18 s | 0.66 | 3.44 | 5.23 | .261 | .298 |
|  |  |  | 15 | 19 s | 0.28 | 1.46 | 5.23 | .392 | .418 |
|  |  |  | 15 | 20 s | -0.10 | -0.50 | 5.23 | .538 | .538 |
| **LITG** | **15** | **1.40** | **15, 210** | **Overall time effect** |  |  |  | **.150** | **.284** |
| **LSMG** | **16** | **0.23** | **15, 225** | **Overall time effect** |  |  |  | **.999** | **.999** |
| **LAG** | **14** | **1.63** | **15, 195** | **Overall time effect** |  |  |  | **.069** | **.167** |
| **RSFG** | **16** | **1.93** | **15, 225** | **Overall time effect** |  |  |  | **.022** | **.061** |
| **RMFG** | **16** | **1.14** | **15, 225** | **Overall time effect** |  |  |  | **.318** | **.541** |
| **RIFGtri** | **16** | **0.87** | **15, 225** | **Overall time effect** |  |  |  | **.600** | **.765** |
| **RIFGop** | **16** | **0.84** | **15, 225** | **Overall time effect** |  |  |  | **.630** | **.765** |
| **RSTG** | **16** | **2.18** | **15, 225** | **Overall time effect** |  |  |  | **.008** | **.033** |
|  |  |  | 15 | 5 s | 0.00 | -0.01 | 3.08 | .502 | .908 |
|  |  |  | 15 | 6 s | 0.16 | 0.49 | 3.08 | .438 | .908 |
|  |  |  | 15 | 7 s | 0.25 | 0.78 | 3.08 | .401 | .908 |
|  |  |  | 15 | 8 s | 0.22 | 0.68 | 3.08 | .415 | .908 |
|  |  |  | 15 | 9 s | 0.12 | 0.36 | 3.08 | .454 | .908 |
|  |  |  | 15 | 10 s | -0.06 | -0.17 | 3.08 | .522 | .908 |
|  |  |  | 15 | 11 s | -0.15 | -0.47 | 3.08 | .560 | .908 |
|  |  |  | 15 | 12 s | -0.17 | -0.54 | 3.08 | .568 | .908 |
|  |  |  | 15 | 13 s | -0.11 | -0.34 | 3.08 | .544 | .908 |
|  |  |  | 15 | 14 s | -0.17 | -0.52 | 3.08 | .565 | .908 |
|  |  |  | 15 | 15 s | -0.32 | -0.99 | 3.08 | .624 | .908 |
|  |  |  | 15 | 16 s | -0.63 | -1.94 | 3.08 | .731 | .975 |
|  |  |  | 15 | 17 s | -1.04 | -3.19 | 3.08 | .842 | .990 |
|  |  |  | 15 | 18 s | -1.51 | -4.64 | 3.08 | .924 | .990 |
|  |  |  | 15 | 19 s | -2.01 | -6.19 | 3.08 | .969 | .990 |
|  |  |  | 15 | 20 s | -2.61 | -8.04 | 3.08 | .990 | .990 |
| **RMTG** | **16** | **1.10** | **15, 225** | **Overall time effect** |  |  |  | **.361** | **.557** |
| **RSMG** | **16** | **0.57** | **15, 225** | **Overall time effect** |  |  |  | **.896** | **.999** |
| **RAG** | **15** | **1.57** | **15, 210** | **Overall time effect** |  |  |  | **.083** | **.176** |
| **HbR** |  |  |  |  |  |  |  |  |  |
| **LSFG** | **16** | **0.80** | **15, 225** | **Overall time effect** |  |  |  | **.677** | **.792** |
| **LMFG** | **16** | **1.19** | **15, 225** | **Overall time effect** |  |  |  | **.284** | **.578** |
| **LIFGtri** | **16** | **3.12** | **15, 225** | **Overall time effect** |  |  |  | **< .001** | **.002** |
|  |  |  | 15 | 5 s | -0.84 | -1.23 | 1.47 | .208 | .208 |
|  |  |  | 15 | 6 s | -1.07 | -1.57 | 1.47 | .151 | .161 |
|  |  |  | 15 | 7 s | -1.37 | -2.00 | 1.47 | .096 | .109 |
|  |  |  | 15 | 8 s | -1.75 | -2.57 | 1.47 | .050 | .062 |
|  |  |  | 15 | 9 s | -2.17 | -3.19 | 1.47 | .023 | .031 |
|  |  |  | 15 | 10 s | -2.61 | -3.82 | 1.47 | .010 | .014 |
|  |  |  | 15 | 11 s | -2.98 | -4.37 | 1.47 | .005 | .007 |
|  |  |  | 15 | 12 s | -3.22 | -4.72 | 1.47 | .003 | .005 |
|  |  |  | 15 | 13 s | -3.32 | -4.86 | 1.47 | .002 | .005 |
|  |  |  | 15 | 14 s | -3.47 | -5.08 | 1.47 | .002 | .004 |
|  |  |  | 15 | 15 s | -3.63 | -5.32 | 1.47 | .001 | .003 |
|  |  |  | 15 | 16 s | -3.74 | -5.47 | 1.47 | < .001 | .003 |
|  |  |  | 15 | 17 s | -3.77 | -5.53 | 1.47 | < .001 | .003 |
|  |  |  | 15 | 18 s | -3.84 | -5.62 | 1.47 | < .001 | .003 |
|  |  |  | 15 | 19 s | -3.83 | -5.62 | 1.47 | < .001 | .003 |
|  |  |  | 15 | 20 s | -3.89 | -5.70 | 1.47 | < .001 | .003 |
| **LIFGop** | **16** | **1.68** | **15, 225** | **Overall time effect** |  |  |  | **.056** | **.240** |
| **LSTG** | **16** | **1.28** | **15, 225** | **Overall time effect** |  |  |  | **.219** | **.532** |
| **LMTG** | **16** | **1.48** | **15, 225** | **Overall time effect** |  |  |  | **.115** | **.390** |
| **LITG** | **15** | **0.30** | **15, 210** | **Overall time effect** |  |  |  | **.995** | **.995** |
| **LSMG** | **16** | **0.78** | **15, 225** | **Overall time effect** |  |  |  | **.699** | **.792** |
| **LAG** | **14** | **0.89** | **15, 195** | **Overall time effect** |  |  |  | **.580** | **.758** |
| **RSFG** | **16** | **1.16** | **15, 225** | **Overall time effect** |  |  |  | **.306** | **.578** |
| **RMFG** | **16** | **0.90** | **15, 225** | **Overall time effect** |  |  |  | **.565** | **.758** |
| **RIFGtri** | **16** | **1.74** | **15, 225** | **Overall time effect** |  |  |  | **.045** | **.240** |
| **RIFGop** | **16** | **0.53** | **15, 225** | **Overall time effect** |  |  |  | **.921** | **.978** |
| **RSTG** | **16** | **1.91** | **15, 225** | **Overall time effect** |  |  |  | **.023** | **.199** |
| **RMTG** | **16** | **0.97** | **15, 225** | **Overall time effect** |  |  |  | **.488** | **.758** |
| **RSMG** | **16** | **1.35** | **15, 225** | **Overall time effect** |  |  |  | **.176** | **.498** |
| **RAG** | **15** | **0.93** | **15, 210** | **Overall time effect** |  |  |  | **.534** | **.758** |

Note. Estimated marginal means (EMM) and standard error (SE) values are reported in µM. Model-level *q*-values reflect correction at the false discovery rate (FDR) across regions of interest (ROIs) within each Group x Task x Hemoglobin analysis set. EMM *q*-values reflect FDR correction across the 16 tested time points (5-20 s) within each ROI. Oxyhemoglobin (HbO) EMMs were tested against 0 in the positive direction; de-oxyhemoglobin (HbR) EMMs were tested against 0 in the negative direction. EMMs are shown only when the model-level FDR-corrected result is significant. AG = angular gyrus, IFGtri/op = inferior frontal gyrus, pars triangularis/opercularis, ITG = inferior temporal gyrus, L = left, MFG = middle frontal gyrus, MTG = middle temporal gyrus, R = right, SFG = superior frontal gyrus, SMG = supramarginal gyrus, STG = superior temporal gyrus.

Supplemental Table 3. Linear mixed effects models results for Semantic Matching for participants with aphasia

| **Region** | ***N*** | ***F*** | ***df*** | **Effect / time** | ***t*** | **EMM (µM)** | **SE (µM)** | ***p*** | ***q* (FDR)** |
| --- | --- | --- | --- | --- | --- | --- | --- | --- | --- |
| **HbO** |  |  |  |  |  |  |  |  |  |
| **LSFG** | **15** | **1.54** | **15, 210** | **Overall time effect** |  |  |  | **.092** | **.522** |
| **LMFG** | **15** | **0.94** | **15, 210** | **Overall time effect** |  |  |  | **.522** | **.999** |
| **LIFGtri** | **13** | **1.30** | **15, 180** | **Overall time effect** |  |  |  | **.205** | **.697** |
| **LIFGop** | **11** | **0.85** | **15, 150** | **Overall time effect** |  |  |  | **.621** | **.999** |
| **LSTG** | **7** | **0.30** | **15, 90** | **Overall time effect** |  |  |  | **.994** | **.999** |
| **LMTG** | **12** | **0.76** | **15, 165** | **Overall time effect** |  |  |  | **.715** | **.999** |
| **LITG** | **12** | **0.70** | **15, 165** | **Overall time effect** |  |  |  | **.785** | **.999** |
| **LSMG** | **12** | **0.37** | **15, 165** | **Overall time effect** |  |  |  | **.984** | **.999** |
| **LAG** | **8** | **0.43** | **15, 105** | **Overall time effect** |  |  |  | **.966** | **.999** |
| **RSFG** | **15** | **1.34** | **15, 210** | **Overall time effect** |  |  |  | **.178** | **.697** |
| **RMFG** | **15** | **2.19** | **15, 210** | **Overall time effect** |  |  |  | **.008** | **.066** |
| **RIFGtri** | **15** | **2.64** | **15, 210** | **Overall time effect** |  |  |  | **.001** | **.019** |
|  |  |  | 14 | 5 s | -1.49 | -4.78 | 3.22 | .920 | .999 |
|  |  |  | 14 | 6 s | -1.53 | -4.91 | 3.22 | .925 | .999 |
|  |  |  | 14 | 7 s | -1.47 | -4.73 | 3.22 | .918 | .999 |
|  |  |  | 14 | 8 s | -1.28 | -4.11 | 3.22 | .889 | .999 |
|  |  |  | 14 | 9 s | -1.03 | -3.32 | 3.22 | .840 | .999 |
|  |  |  | 14 | 10 s | -0.86 | -2.78 | 3.22 | .799 | .999 |
|  |  |  | 14 | 11 s | -0.79 | -2.55 | 3.22 | .779 | .999 |
|  |  |  | 14 | 12 s | -0.89 | -2.85 | 3.22 | .805 | .999 |
|  |  |  | 14 | 13 s | -1.21 | -3.90 | 3.22 | .877 | .999 |
|  |  |  | 14 | 14 s | -1.70 | -5.46 | 3.22 | .944 | .999 |
|  |  |  | 14 | 15 s | -2.27 | -7.31 | 3.22 | .980 | .999 |
|  |  |  | 14 | 16 s | -2.87 | -9.25 | 3.22 | .994 | .999 |
|  |  |  | 14 | 17 s | -3.34 | -10.75 | 3.22 | .998 | .999 |
|  |  |  | 14 | 18 s | -3.60 | -11.58 | 3.22 | .999 | .999 |
|  |  |  | 14 | 19 s | -3.57 | -11.48 | 3.22 | .998 | .999 |
|  |  |  | 14 | 20 s | -3.38 | -10.88 | 3.22 | .998 | .999 |
| **RIFGop** | **15** | **1.08** | **15, 210** | **Overall time effect** |  |  |  | **.380** | **.999** |
| **RSTG** | **15** | **0.25** | **15, 210** | **Overall time effect** |  |  |  | **.998** | **.999** |
| **RMTG** | **15** | **0.52** | **15, 210** | **Overall time effect** |  |  |  | **.927** | **.999** |
| **RSMG** | **15** | **0.28** | **15, 210** | **Overall time effect** |  |  |  | **.996** | **.999** |
| **RAG** | **15** | **0.24** | **15, 210** | **Overall time effect** |  |  |  | **.999** | **.999** |
| **HbR** |  |  |  |  |  |  |  |  |  |
| **LSFG** | **15** | **1.85** | **15, 210** | **Overall time effect** |  |  |  | **.029** | **.100** |
| **LMFG** | **15** | **4.12** | **15, 210** | **Overall time effect** |  |  |  | **< .001** | **< .001** |
|  |  |  | 14 | 5 s | -0.49 | -0.48 | 0.99 | .316 | .794 |
|  |  |  | 14 | 6 s | -0.52 | -0.51 | 0.99 | .305 | .794 |
|  |  |  | 14 | 7 s | -0.51 | -0.50 | 0.99 | .309 | .794 |
|  |  |  | 14 | 8 s | -0.47 | -0.47 | 0.99 | .322 | .794 |
|  |  |  | 14 | 9 s | -0.39 | -0.38 | 0.99 | .352 | .794 |
|  |  |  | 14 | 10 s | -0.25 | -0.25 | 0.99 | .403 | .794 |
|  |  |  | 14 | 11 s | -0.10 | -0.10 | 0.99 | .462 | .794 |
|  |  |  | 14 | 12 s | -0.03 | -0.03 | 0.99 | .487 | .794 |
|  |  |  | 14 | 13 s | -0.05 | -0.05 | 0.99 | .479 | .794 |
|  |  |  | 14 | 14 s | 0.01 | 0.01 | 0.99 | .505 | .794 |
|  |  |  | 14 | 15 s | 0.12 | 0.12 | 0.99 | .546 | .794 |
|  |  |  | 14 | 16 s | 0.34 | 0.33 | 0.99 | .629 | .839 |
|  |  |  | 14 | 17 s | 0.74 | 0.73 | 0.99 | .765 | .941 |
|  |  |  | 14 | 18 s | 1.32 | 1.31 | 0.99 | .897 | .987 |
|  |  |  | 14 | 19 s | 1.94 | 1.92 | 0.99 | .964 | .987 |
|  |  |  | 14 | 20 s | 2.50 | 2.46 | 0.99 | .987 | .987 |
| **LIFGtri** | **13** | **2.10** | **15, 180** | **Overall time effect** |  |  |  | **.012** | **.049** |
|  |  |  | 12 | 5 s | -2.46 | -2.50 | 1.02 | .015 | .015 |
|  |  |  | 12 | 6 s | -3.03 | -3.09 | 1.02 | .005 | .006 |
|  |  |  | 12 | 7 s | -3.52 | -3.58 | 1.02 | .002 | .002 |
|  |  |  | 12 | 8 s | -3.80 | -3.86 | 1.02 | .001 | .002 |
|  |  |  | 12 | 9 s | -3.99 | -4.05 | 1.02 | < .001 | .001 |
|  |  |  | 12 | 10 s | -4.17 | -4.24 | 1.02 | < .001 | .001 |
|  |  |  | 12 | 11 s | -4.27 | -4.34 | 1.02 | < .001 | .001 |
|  |  |  | 12 | 12 s | -4.41 | -4.48 | 1.02 | < .001 | .001 |
|  |  |  | 12 | 13 s | -4.67 | -4.75 | 1.02 | < .001 | < .001 |
|  |  |  | 12 | 14 s | -4.87 | -4.96 | 1.02 | < .001 | < .001 |
|  |  |  | 12 | 15 s | -4.91 | -4.99 | 1.02 | < .001 | < .001 |
|  |  |  | 12 | 16 s | -4.90 | -4.98 | 1.02 | < .001 | < .001 |
|  |  |  | 12 | 17 s | -4.67 | -4.75 | 1.02 | < .001 | < .001 |
|  |  |  | 12 | 18 s | -4.39 | -4.47 | 1.02 | < .001 | .001 |
|  |  |  | 12 | 19 s | -4.20 | -4.27 | 1.02 | < .001 | .001 |
|  |  |  | 12 | 20 s | -4.07 | -4.14 | 1.02 | < .001 | .001 |
| **LIFGop** | **11** | **0.20** | **15, 150** | **Overall time effect** |  |  |  | **.999** | **.999** |
| **LSTG** | **7** | **0.52** | **15, 90** | **Overall time effect** |  |  |  | **.925** | **.999** |
| **LMTG** | **12** | **1.40** | **15, 165** | **Overall time effect** |  |  |  | **.155** | **.403** |
| **LITG** | **12** | **2.17** | **15, 165** | **Overall time effect** |  |  |  | **.009** | **.049** |
|  |  |  | 11 | 5 s | -0.95 | -0.92 | 0.97 | .181 | .207 |
|  |  |  | 11 | 6 s | -1.30 | -1.26 | 0.97 | .110 | .147 |
|  |  |  | 11 | 7 s | -1.55 | -1.50 | 0.97 | .075 | .109 |
|  |  |  | 11 | 8 s | -1.98 | -1.91 | 0.97 | .037 | .065 |
|  |  |  | 11 | 9 s | -2.47 | -2.39 | 0.97 | .015 | .031 |
|  |  |  | 11 | 10 s | -2.91 | -2.82 | 0.97 | .007 | .019 |
|  |  |  | 11 | 11 s | -3.30 | -3.19 | 0.97 | .004 | .012 |
|  |  |  | 11 | 12 s | -3.62 | -3.50 | 0.97 | .002 | .011 |
|  |  |  | 11 | 13 s | -3.74 | -3.62 | 0.97 | .002 | .011 |
|  |  |  | 11 | 14 s | -3.58 | -3.47 | 0.97 | .002 | .011 |
|  |  |  | 11 | 15 s | -3.25 | -3.14 | 0.97 | .004 | .012 |
|  |  |  | 11 | 16 s | -2.70 | -2.62 | 0.97 | .010 | .023 |
|  |  |  | 11 | 17 s | -1.91 | -1.85 | 0.97 | .041 | .066 |
|  |  |  | 11 | 18 s | -1.10 | -1.06 | 0.97 | .148 | .182 |
|  |  |  | 11 | 19 s | -0.38 | -0.37 | 0.97 | .354 | .378 |
|  |  |  | 11 | 20 s | 0.19 | 0.18 | 0.97 | .572 | .572 |
| **LSMG** | **12** | **1.07** | **15, 165** | **Overall time effect** |  |  |  | **.386** | **.671** |
| **LAG** | **8** | **0.49** | **15, 105** | **Overall time effect** |  |  |  | **.940** | **.999** |
| **RSFG** | **15** | **1.06** | **15, 210** | **Overall time effect** |  |  |  | **.395** | **.671** |
| **RMFG** | **15** | **3.11** | **15, 210** | **Overall time effect** |  |  |  | **< .001** | **.001** |
|  |  |  | 14 | 5 s | 0.49 | 0.70 | 1.42 | .685 | .998 |
|  |  |  | 14 | 6 s | 0.75 | 1.06 | 1.42 | .766 | .998 |
|  |  |  | 14 | 7 s | 0.95 | 1.35 | 1.42 | .821 | .998 |
|  |  |  | 14 | 8 s | 1.18 | 1.66 | 1.42 | .870 | .998 |
|  |  |  | 14 | 9 s | 1.41 | 1.99 | 1.42 | .909 | .998 |
|  |  |  | 14 | 10 s | 1.61 | 2.28 | 1.42 | .936 | .998 |
|  |  |  | 14 | 11 s | 1.79 | 2.53 | 1.42 | .952 | .998 |
|  |  |  | 14 | 12 s | 1.90 | 2.68 | 1.42 | .961 | .998 |
|  |  |  | 14 | 13 s | 1.92 | 2.72 | 1.42 | .962 | .998 |
|  |  |  | 14 | 14 s | 1.97 | 2.79 | 1.42 | .966 | .998 |
|  |  |  | 14 | 15 s | 2.02 | 2.87 | 1.42 | .969 | .998 |
|  |  |  | 14 | 16 s | 2.15 | 3.05 | 1.42 | .975 | .998 |
|  |  |  | 14 | 17 s | 2.38 | 3.37 | 1.42 | .984 | .998 |
|  |  |  | 14 | 18 s | 2.73 | 3.86 | 1.42 | .992 | .998 |
|  |  |  | 14 | 19 s | 3.05 | 4.31 | 1.42 | .996 | .998 |
|  |  |  | 14 | 20 s | 3.36 | 4.76 | 1.42 | .998 | .998 |
| **RIFGtri** | **15** | **1.37** | **15, 210** | **Overall time effect** |  |  |  | **.166** | **.403** |
| **RIFGop** | **15** | **0.58** | **15, 210** | **Overall time effect** |  |  |  | **.892** | **.999** |
| **RSTG** | **15** | **0.26** | **15, 210** | **Overall time effect** |  |  |  | **.998** | **.999** |
| **RMTG** | **15** | **0.68** | **15, 210** | **Overall time effect** |  |  |  | **.802** | **.999** |
| **RSMG** | **15** | **0.74** | **15, 210** | **Overall time effect** |  |  |  | **.746** | **.999** |
| **RAG** | **15** | **1.09** | **15, 210** | **Overall time effect** |  |  |  | **.368** | **.671** |

Note. Estimated marginal means (EMM) and standard error (SE) values are reported in µM. Model-level *q*-values reflect correction at the false discovery rate (FDR) across regions of interest (ROIs) within each Group x Task x Hemoglobin analysis set. EMM *q*-values reflect FDR correction across the 16 tested time points (5-20 s) within each ROI. Oxyhemoglobin (HbO) EMMs were tested against 0 in the positive direction; de-oxyhemoglobin (HbR) EMMs were tested against 0 in the negative direction. EMMs are shown only when the model-level FDR-corrected result is significant. AG = angular gyrus, IFGtri/op = inferior frontal gyrus, pars triangularis/opercularis, ITG = inferior temporal gyrus, L = left, MFG = middle frontal gyrus, MTG = middle temporal gyrus, R = right, SFG = superior frontal gyrus, SMG = supramarginal gyrus, STG = superior temporal gyrus.

Supplemental Table 4. Linear mixed effects models comparing Semantic Matching results between PWA and healthy controls

| **Region** | ***N* (PWA /**  **Controls)** | ***F*** | ***df*** | **Effect** | ***t*** | **PWA -**  **Controls (µM)** | ***SE* (µM)** | ***p*** | ***q* (FDR)** |
| --- | --- | --- | --- | --- | --- | --- | --- | --- | --- |
| **HbO** | | | | | | | | | |
| LSFG | 15 / 16 | 0.01 | 1, 29 | Group |  |  |  | .939 | .987 |
| LSFG | 15 / 16 | 1.73 | 15, 435 | Time |  |  |  | .043 | .238 |
| LSFG | 15 / 16 | 1.40 | 15, 435 | Group x Time |  |  |  | .145 | .308 |
| LMFG | 15 / 16 | 0.63 | 1, 29 | Group |  |  |  | .435 | .987 |
| LMFG | 15 / 16 | 1.09 | 15, 435 | Time |  |  |  | .359 | .758 |
| LMFG | 15 / 16 | 0.89 | 15, 435 | Group x Time |  |  |  | .578 | .819 |
| LIFGtri | 13 / 16 | 0.92 | 1, 27 | Group |  |  |  | .347 | .987 |
| LIFGtri | 13 / 16 | 1.47 | 15, 405 | Time |  |  |  | .112 | .380 |
| LIFGtri | 13 / 16 | 1.77 | 15, 405 | Group x Time |  |  |  | .037 | .156 |
| LIFGop | 11 / 16 | 0.05 | 1, 25 | Group |  |  |  | .831 | .987 |
| LIFGop | 11 / 16 | 1.16 | 15, 375 | Time |  |  |  | .300 | .729 |
| LIFGop | 11 / 16 | 2.43 | 15, 375 | Group x Time |  |  |  | .002 | .037 |
|  |  |  | 25 | 5 s | -0.22 | -1.64 | 7.59 | .831 | .831 |
|  |  |  | 25 | 6 s | -0.52 | -3.93 | 7.59 | .609 | .650 |
|  |  |  | 25 | 7 s | -0.76 | -5.80 | 7.59 | .452 | .516 |
|  |  |  | 25 | 8 s | -0.98 | -7.41 | 7.59 | .338 | .416 |
|  |  |  | 25 | 9 s | -1.16 | -8.84 | 7.59 | .255 | .371 |
|  |  |  | 25 | 10 s | -1.29 | -9.82 | 7.59 | .208 | .332 |
|  |  |  | 25 | 11 s | -1.49 | -11.35 | 7.59 | .148 | .262 |
|  |  |  | 25 | 12 s | -1.91 | -14.52 | 7.59 | .067 | .154 |
|  |  |  | 25 | 13 s | -2.43 | -18.45 | 7.59 | .023 | .069 |
|  |  |  | 25 | 14 s | -2.78 | -21.14 | 7.59 | .010 | .040 |
|  |  |  | 25 | 15 s | -2.95 | -22.40 | 7.59 | .007 | .040 |
|  |  |  | 25 | 16 s | -2.95 | -22.42 | 7.59 | .007 | .040 |
|  |  |  | 25 | 17 s | -2.81 | -21.34 | 7.59 | .009 | .040 |
|  |  |  | 25 | 18 s | -2.37 | -17.99 | 7.59 | .026 | .069 |
|  |  |  | 25 | 19 s | -1.78 | -13.49 | 7.59 | .088 | .175 |
|  |  |  | 25 | 20 s | -1.10 | -8.36 | 7.59 | .281 | .375 |
| LSTG | 7 / 16 | 0.02 | 1, 21 | Group |  |  |  | .896 | .987 |
| LSTG | 7 / 16 | 0.43 | 15, 315 | Time |  |  |  | .968 | .990 |
| LSTG | 7 / 16 | 1.43 | 15, 315 | Group x Time |  |  |  | .132 | .308 |
| LMTG | 12 / 16 | 0.00 | 1, 26 | Group |  |  |  | .987 | .987 |
| LMTG | 12 / 16 | 0.96 | 15, 390 | Time |  |  |  | .503 | .777 |
| LMTG | 12 / 16 | 1.07 | 15, 390 | Group x Time |  |  |  | .385 | .627 |
| LITG | 12 / 15 | 0.53 | 1, 25 | Group |  |  |  | .473 | .987 |
| LITG | 12 / 15 | 1.05 | 15, 375 | Time |  |  |  | .401 | .758 |
| LITG | 12 / 15 | 1.67 | 15, 375 | Group x Time |  |  |  | .055 | .188 |
| LSMG | 12 / 16 | 0.02 | 1, 26 | Group |  |  |  | .878 | .987 |
| LSMG | 12 / 16 | 0.73 | 15, 390 | Time |  |  |  | .758 | .990 |
| LSMG | 12 / 16 | 0.59 | 15, 390 | Group x Time |  |  |  | .882 | .937 |
| LAG | 8 / 14 | 0.30 | 1, 20 | Group |  |  |  | .590 | .987 |
| LAG | 8 / 14 | 0.96 | 15, 300 | Time |  |  |  | .499 | .777 |
| LAG | 8 / 14 | 1.27 | 15, 300 | Group x Time |  |  |  | .219 | .413 |
| RSFG | 15 / 16 | 0.34 | 1, 29 | Group |  |  |  | .567 | .987 |
| RSFG | 15 / 16 | 1.66 | 15, 435 | Time |  |  |  | .056 | .238 |
| RSFG | 15 / 16 | 1.56 | 15, 435 | Group x Time |  |  |  | .080 | .227 |
| RMFG | 15 / 16 | 0.01 | 1, 29 | Group |  |  |  | .913 | .987 |
| RMFG | 15 / 16 | 2.54 | 15, 435 | Time |  |  |  | .001 | .021 |
| RMFG | 15 / 16 | 1.89 | 15, 435 | Group x Time |  |  |  | .022 | .126 |
| RIFGtri | 15 / 16 | 0.20 | 1, 29 | Group |  |  |  | .661 | .987 |
| RIFGtri | 15 / 16 | 2.32 | 15, 435 | Time |  |  |  | .003 | .029 |
| RIFGtri | 15 / 16 | 1.96 | 15, 435 | Group x Time |  |  |  | .017 | .126 |
| RIFGop | 15 / 16 | 0.76 | 1, 29 | Group |  |  |  | .391 | .987 |
| RIFGop | 15 / 16 | 1.38 | 15, 435 | Time |  |  |  | .155 | .439 |
| RIFGop | 15 / 16 | 0.61 | 15, 435 | Group x Time |  |  |  | .864 | .937 |
| RSTG | 15 / 16 | 1.12 | 1, 29 | Group |  |  |  | .299 | .987 |
| RSTG | 15 / 16 | 0.34 | 15, 435 | Time |  |  |  | .990 | .990 |
| RSTG | 15 / 16 | 1.05 | 15, 435 | Group x Time |  |  |  | .405 | .627 |
| RMTG | 15 / 16 | 2.37 | 1, 29 | Group |  |  |  | .135 | .987 |
| RMTG | 15 / 16 | 0.70 | 15, 435 | Time |  |  |  | .787 | .990 |
| RMTG | 15 / 16 | 0.82 | 15, 435 | Group x Time |  |  |  | .657 | .851 |
| RSMG | 15 / 16 | 0.01 | 1, 29 | Group |  |  |  | .915 | .987 |
| RSMG | 15 / 16 | 0.41 | 15, 435 | Time |  |  |  | .975 | .990 |
| RSMG | 15 / 16 | 0.47 | 15, 435 | Group x Time |  |  |  | .957 | .957 |
| RAG | 15 / 15 | 0.09 | 1, 28 | Group |  |  |  | .770 | .987 |
| RAG | 15 / 15 | 0.37 | 15, 420 | Time |  |  |  | .987 | .990 |
| RAG | 15 / 15 | 0.78 | 15, 420 | Group x Time |  |  |  | .700 | .851 |
| **HbR** | | | | | | | | | |
| LSFG | 15 / 16 | 0.09 | 1, 29 | Group |  |  |  | .764 | .944 |
| LSFG | 15 / 16 | 2.58 | 15, 435 | Time |  |  |  | .001 | .006 |
| LSFG | 15 / 16 | 0.88 | 15, 435 | Group x Time |  |  |  | .583 | .901 |
| LMFG | 15 / 16 | 0.05 | 1, 29 | Group |  |  |  | .820 | .944 |
| LMFG | 15 / 16 | 4.39 | 15, 435 | Time |  |  |  | < .001 | < .001 |
| LMFG | 15 / 16 | 2.20 | 15, 435 | Group x Time |  |  |  | .006 | .100 |
| LIFGtri | 13 / 16 | 0.47 | 1, 27 | Group |  |  |  | .500 | .944 |
| LIFGtri | 13 / 16 | 1.57 | 15, 405 | Time |  |  |  | .078 | .191 |
| LIFGtri | 13 / 16 | 0.96 | 15, 405 | Group x Time |  |  |  | .499 | .848 |
| LIFGop | 11 / 16 | 0.16 | 1, 25 | Group |  |  |  | .695 | .944 |
| LIFGop | 11 / 16 | 0.11 | 15, 375 | Time |  |  |  | 1.000 | 1.000 |
| LIFGop | 11 / 16 | 1.21 | 15, 375 | Group x Time |  |  |  | .262 | .636 |
| LSTG | 7 / 16 | 0.01 | 1, 21 | Group |  |  |  | .915 | .944 |
| LSTG | 7 / 16 | 0.33 | 15, 315 | Time |  |  |  | .991 | 1.000 |
| LSTG | 7 / 16 | 0.56 | 15, 315 | Group x Time |  |  |  | .906 | .974 |
| LMTG | 12 / 16 | 0.27 | 1, 26 | Group |  |  |  | .607 | .944 |
| LMTG | 12 / 16 | 0.98 | 15, 390 | Time |  |  |  | .477 | .737 |
| LMTG | 12 / 16 | 0.70 | 15, 390 | Group x Time |  |  |  | .790 | .974 |
| LITG | 12 / 15 | 0.10 | 1, 25 | Group |  |  |  | .753 | .944 |
| LITG | 12 / 15 | 2.34 | 15, 375 | Time |  |  |  | .003 | .014 |
| LITG | 12 / 15 | 1.10 | 15, 375 | Group x Time |  |  |  | .358 | .711 |
| LSMG | 12 / 16 | 0.44 | 1, 26 | Group |  |  |  | .514 | .944 |
| LSMG | 12 / 16 | 1.80 | 15, 390 | Time |  |  |  | .033 | .094 |
| LSMG | 12 / 16 | 1.50 | 15, 390 | Group x Time |  |  |  | .101 | .575 |
| LAG | 8 / 14 | 0.24 | 1, 20 | Group |  |  |  | .630 | .944 |
| LAG | 8 / 14 | 1.21 | 15, 300 | Time |  |  |  | .265 | .470 |
| LAG | 8 / 14 | 0.80 | 15, 300 | Group x Time |  |  |  | .675 | .956 |
| RSFG | 15 / 16 | 0.01 | 1, 29 | Group |  |  |  | .933 | .944 |
| RSFG | 15 / 16 | 1.26 | 15, 435 | Time |  |  |  | .222 | .470 |
| RSFG | 15 / 16 | 0.51 | 15, 435 | Group x Time |  |  |  | .937 | .974 |
| RMFG | 15 / 16 | 0.01 | 1, 29 | Group |  |  |  | .944 | .944 |
| RMFG | 15 / 16 | 3.62 | 15, 435 | Time |  |  |  | < .001 | < .001 |
| RMFG | 15 / 16 | 1.70 | 15, 435 | Group x Time |  |  |  | .049 | .415 |
| RIFGtri | 15 / 16 | 0.69 | 1, 29 | Group |  |  |  | .412 | .944 |
| RIFGtri | 15 / 16 | 1.19 | 15, 435 | Time |  |  |  | .276 | .470 |
| RIFGtri | 15 / 16 | 1.29 | 15, 435 | Group x Time |  |  |  | .205 | .636 |
| RIFGop | 15 / 16 | 1.16 | 1, 29 | Group |  |  |  | .289 | .944 |
| RIFGop | 15 / 16 | 0.71 | 15, 435 | Time |  |  |  | .771 | 1.000 |
| RIFGop | 15 / 16 | 0.42 | 15, 435 | Group x Time |  |  |  | .974 | .974 |
| RSTG | 15 / 16 | 0.01 | 1, 29 | Group |  |  |  | .939 | .944 |
| RSTG | 15 / 16 | 0.15 | 15, 435 | Time |  |  |  | 1.000 | 1.000 |
| RSTG | 15 / 16 | 1.26 | 15, 435 | Group x Time |  |  |  | .226 | .636 |
| RMTG | 15 / 16 | 0.07 | 1, 29 | Group |  |  |  | .797 | .944 |
| RMTG | 15 / 16 | 0.57 | 15, 435 | Time |  |  |  | .898 | 1.000 |
| RMTG | 15 / 16 | 1.08 | 15, 435 | Group x Time |  |  |  | .376 | .711 |
| RSMG | 15 / 16 | 2.82 | 1, 29 | Group |  |  |  | .104 | .944 |
| RSMG | 15 / 16 | 0.31 | 15, 435 | Time |  |  |  | .995 | 1.000 |
| RSMG | 15 / 16 | 1.33 | 15, 435 | Group x Time |  |  |  | .183 | .636 |
| RAG | 15 / 15 | 0.11 | 1, 28 | Group |  |  |  | .747 | .944 |
| RAG | 15 / 15 | 1.80 | 15, 420 | Time |  |  |  | .032 | .094 |
| RAG | 15 / 15 | 0.68 | 15, 420 | Group x Time |  |  |  | .809 | .974 |

Note. Differences between PWA and controls in estimated marginal means and standard error (SE) values are reported in µM. Model-level *q*-values reflect correction at the false discovery rate (FDR) across regions of interest (ROIs) within each Group x Task x Hemoglobin analysis set. EMM *q*-values reflect FDR correction across the 16 tested time points (5-20 s) within each ROI. Oxyhemoglobin (HbO) EMMs were tested against 0 in the positive direction; de-oxyhemoglobin (HbR) EMMs were tested against 0 in the negative direction. EMMs are shown only when the model-level FDR-corrected result for the Group x Time interaction is significant. AG = angular gyrus, IFGtri/op = inferior frontal gyrus, pars triangularis/opercularis, ITG = inferior temporal gyrus, L = left, MFG = middle frontal gyrus, MTG = middle temporal gyrus, R = right, SFG = superior frontal gyrus, SMG = supramarginal gyrus, STG = superior temporal gyrus.

Supplemental Table 5. Linear mixed effects models results for Rhyme Judgment for healthy controls

| **Region** | ***N*** | ***F*** | ***df*** | **Effect / time** | ***t*** | **EMM (µM)** | **SE (µM)** | ***p*** | ***q* (FDR)** |
| --- | --- | --- | --- | --- | --- | --- | --- | --- | --- |
| **HbO** |  |  |  |  |  |  |  |  |  |
| **LSFG** | **16** | **0.91** | **15, 225** | **Overall time effect** |  |  |  | **.555** | **.786** |
| **LMFG** | **16** | **1.19** | **15, 225** | **Overall time effect** |  |  |  | **.277** | **.470** |
| **LIFGtri** | **16** | **2.34** | **15, 225** | **Overall time effect** |  |  |  | **.004** | **.020** |
|  |  |  | 15 | 5 s | 1.46 | 2.55 | 1.74 | .082 | .095 |
|  |  |  | 15 | 6 s | 1.43 | 2.49 | 1.74 | .087 | .095 |
|  |  |  | 15 | 7 s | 1.52 | 2.65 | 1.74 | .075 | .095 |
|  |  |  | 15 | 8 s | 1.81 | 3.16 | 1.74 | .045 | .066 |
|  |  |  | 15 | 9 s | 2.27 | 3.96 | 1.74 | .019 | .038 |
|  |  |  | 15 | 10 s | 2.68 | 4.66 | 1.74 | .009 | .023 |
|  |  |  | 15 | 11 s | 3.01 | 5.25 | 1.74 | .004 | .019 |
|  |  |  | 15 | 12 s | 3.24 | 5.64 | 1.74 | .003 | .019 |
|  |  |  | 15 | 13 s | 3.24 | 5.65 | 1.74 | .003 | .019 |
|  |  |  | 15 | 14 s | 2.98 | 5.19 | 1.74 | .005 | .019 |
|  |  |  | 15 | 15 s | 2.68 | 4.66 | 1.74 | .009 | .023 |
|  |  |  | 15 | 16 s | 2.37 | 4.13 | 1.74 | .016 | .036 |
|  |  |  | 15 | 17 s | 2.15 | 3.74 | 1.74 | .024 | .043 |
|  |  |  | 15 | 18 s | 1.90 | 3.31 | 1.74 | .038 | .061 |
|  |  |  | 15 | 19 s | 1.41 | 2.46 | 1.74 | .089 | .095 |
|  |  |  | 15 | 20 s | 0.68 | 1.19 | 1.74 | .253 | .253 |
| **LIFGop** | **16** | **2.30** | **15, 225** | **Overall time effect** |  |  |  | **.005** | **.020** |
|  |  |  | 15 | 5 s | 0.28 | 1.40 | 4.98 | .391 | .675 |
|  |  |  | 15 | 6 s | 0.16 | 0.81 | 4.98 | .436 | .675 |
|  |  |  | 15 | 7 s | 0.17 | 0.82 | 4.98 | .436 | .675 |
|  |  |  | 15 | 8 s | 0.28 | 1.39 | 4.98 | .392 | .675 |
|  |  |  | 15 | 9 s | 0.49 | 2.44 | 4.98 | .316 | .675 |
|  |  |  | 15 | 10 s | 0.73 | 3.66 | 4.98 | .237 | .675 |
|  |  |  | 15 | 11 s | 0.92 | 4.60 | 4.98 | .186 | .675 |
|  |  |  | 15 | 12 s | 0.92 | 4.57 | 4.98 | .187 | .675 |
|  |  |  | 15 | 13 s | 0.76 | 3.80 | 4.98 | .229 | .675 |
|  |  |  | 15 | 14 s | 0.44 | 2.20 | 4.98 | .333 | .675 |
|  |  |  | 15 | 15 s | 0.09 | 0.46 | 4.98 | .464 | .675 |
|  |  |  | 15 | 16 s | -0.20 | -0.98 | 4.98 | .577 | .769 |
|  |  |  | 15 | 17 s | -0.41 | -2.03 | 4.98 | .655 | .806 |
|  |  |  | 15 | 18 s | -0.68 | -3.37 | 4.98 | .745 | .852 |
|  |  |  | 15 | 19 s | -0.97 | -4.82 | 4.98 | .825 | .881 |
|  |  |  | 15 | 20 s | -1.35 | -6.73 | 4.98 | .902 | .902 |
| **LSTG** | **16** | **0.39** | **15, 225** | **Overall time effect** |  |  |  | **.982** | **.982** |
| **LMTG** | **16** | **1.01** | **15, 225** | **Overall time effect** |  |  |  | **.448** | **.692** |
| **LITG** | **15** | **1.27** | **15, 210** | **Overall time effect** |  |  |  | **.226** | **.426** |
| **LSMG** | **16** | **3.61** | **15, 225** | **Overall time effect** |  |  |  | **< .001** | **< .001** |
|  |  |  | 15 | 5 s | -0.49 | -1.55 | 3.16 | .684 | .684 |
|  |  |  | 15 | 6 s | -0.09 | -0.28 | 3.16 | .534 | .611 |
|  |  |  | 15 | 7 s | 0.50 | 1.57 | 3.16 | .314 | .456 |
|  |  |  | 15 | 8 s | 1.07 | 3.39 | 3.16 | .150 | .301 |
|  |  |  | 15 | 9 s | 1.63 | 5.16 | 3.16 | .062 | .174 |
|  |  |  | 15 | 10 s | 2.00 | 6.33 | 3.16 | .032 | .143 |
|  |  |  | 15 | 11 s | 2.14 | 6.78 | 3.16 | .025 | .143 |
|  |  |  | 15 | 12 s | 2.12 | 6.71 | 3.16 | .026 | .143 |
|  |  |  | 15 | 13 s | 1.94 | 6.14 | 3.16 | .036 | .143 |
|  |  |  | 15 | 14 s | 1.60 | 5.06 | 3.16 | .065 | .174 |
|  |  |  | 15 | 15 s | 1.17 | 3.71 | 3.16 | .129 | .296 |
|  |  |  | 15 | 16 s | 0.82 | 2.59 | 3.16 | .213 | .379 |
|  |  |  | 15 | 17 s | 0.51 | 1.62 | 3.16 | .308 | .456 |
|  |  |  | 15 | 18 s | 0.22 | 0.70 | 3.16 | .414 | .552 |
|  |  |  | 15 | 19 s | -0.07 | -0.21 | 3.16 | .527 | .611 |
|  |  |  | 15 | 20 s | -0.30 | -0.95 | 3.16 | .616 | .657 |
| **LAG** | **14** | **2.34** | **15, 195** | **Overall time effect** |  |  |  | **.004** | **.020** |
|  |  |  | 13 | 5 s | -1.31 | -3.45 | 2.64 | .894 | .894 |
|  |  |  | 13 | 6 s | -0.87 | -2.30 | 2.64 | .801 | .854 |
|  |  |  | 13 | 7 s | -0.14 | -0.38 | 2.64 | .556 | .635 |
|  |  |  | 13 | 8 s | 0.52 | 1.36 | 2.64 | .307 | .410 |
|  |  |  | 13 | 9 s | 1.09 | 2.86 | 2.64 | .148 | .238 |
|  |  |  | 13 | 10 s | 1.44 | 3.80 | 2.64 | .086 | .173 |
|  |  |  | 13 | 11 s | 1.64 | 4.33 | 2.64 | .062 | .173 |
|  |  |  | 13 | 12 s | 1.80 | 4.75 | 2.64 | .048 | .173 |
|  |  |  | 13 | 13 s | 1.91 | 5.02 | 2.64 | .040 | .173 |
|  |  |  | 13 | 14 s | 1.89 | 4.97 | 2.64 | .041 | .173 |
|  |  |  | 13 | 15 s | 1.70 | 4.48 | 2.64 | .057 | .173 |
|  |  |  | 13 | 16 s | 1.61 | 4.23 | 2.64 | .066 | .173 |
|  |  |  | 13 | 17 s | 1.45 | 3.82 | 2.64 | .086 | .173 |
|  |  |  | 13 | 18 s | 1.10 | 2.89 | 2.64 | .147 | .238 |
|  |  |  | 13 | 19 s | 0.61 | 1.60 | 2.64 | .277 | .402 |
|  |  |  | 13 | 20 s | 0.14 | 0.36 | 2.64 | .447 | .550 |
| **RSFG** | **16** | **0.71** | **15, 225** | **Overall time effect** |  |  |  | **.773** | **.822** |
| **RMFG** | **16** | **0.74** | **15, 225** | **Overall time effect** |  |  |  | **.744** | **.822** |
| **RIFGtri** | **16** | **1.40** | **15, 225** | **Overall time effect** |  |  |  | **.149** | **.317** |
| **RIFGop** | **16** | **0.78** | **15, 225** | **Overall time effect** |  |  |  | **.703** | **.822** |
| **RSTG** | **16** | **1.59** | **15, 225** | **Overall time effect** |  |  |  | **.077** | **.263** |
| **RMTG** | **16** | **1.54** | **15, 225** | **Overall time effect** |  |  |  | **.094** | **.266** |
| **RSMG** | **14** | **1.44** | **15, 195** | **Overall time effect** |  |  |  | **.130** | **.317** |
| **RAG** | **15** | **0.84** | **15, 210** | **Overall time effect** |  |  |  | **.629** | **.822** |
| **HbR** |  |  |  |  |  |  |  |  |  |
| **LSFG** | **16** | **1.42** | **15, 225** | **Overall time effect** |  |  |  | **.139** | **.197** |
| **LMFG** | **16** | **3.37** | **15, 225** | **Overall time effect** |  |  |  | **< .001** | **< .001** |
|  |  |  | 15 | 5 s | 0.18 | 0.15 | 0.83 | .570 | .570 |
|  |  |  | 15 | 6 s | -0.17 | -0.14 | 0.83 | .435 | .464 |
|  |  |  | 15 | 7 s | -0.65 | -0.54 | 0.83 | .262 | .299 |
|  |  |  | 15 | 8 s | -1.14 | -0.95 | 0.83 | .136 | .167 |
|  |  |  | 15 | 9 s | -1.51 | -1.25 | 0.83 | .077 | .102 |
|  |  |  | 15 | 10 s | -1.90 | -1.58 | 0.83 | .039 | .056 |
|  |  |  | 15 | 11 s | -2.26 | -1.88 | 0.83 | .020 | .039 |
|  |  |  | 15 | 12 s | -2.56 | -2.13 | 0.83 | .011 | .034 |
|  |  |  | 15 | 13 s | -2.75 | -2.29 | 0.83 | .007 | .034 |
|  |  |  | 15 | 14 s | -2.83 | -2.35 | 0.83 | .006 | .034 |
|  |  |  | 15 | 15 s | -2.73 | -2.27 | 0.83 | .008 | .034 |
|  |  |  | 15 | 16 s | -2.62 | -2.18 | 0.83 | .010 | .034 |
|  |  |  | 15 | 17 s | -2.47 | -2.06 | 0.83 | .013 | .034 |
|  |  |  | 15 | 18 s | -2.31 | -1.92 | 0.83 | .018 | .039 |
|  |  |  | 15 | 19 s | -2.13 | -1.77 | 0.83 | .025 | .045 |
|  |  |  | 15 | 20 s | -2.06 | -1.72 | 0.83 | .028 | .046 |
| **LIFGtri** | **16** | **4.88** | **15, 225** | **Overall time effect** |  |  |  | **< .001** | **< .001** |
|  |  |  | 15 | 5 s | -0.93 | -1.37 | 1.48 | .185 | .185 |
|  |  |  | 15 | 6 s | -1.39 | -2.06 | 1.48 | .092 | .098 |
|  |  |  | 15 | 7 s | -1.89 | -2.79 | 1.48 | .039 | .045 |
|  |  |  | 15 | 8 s | -2.33 | -3.44 | 1.48 | .017 | .021 |
|  |  |  | 15 | 9 s | -2.68 | -3.96 | 1.48 | .009 | .011 |
|  |  |  | 15 | 10 s | -3.07 | -4.54 | 1.48 | .004 | .006 |
|  |  |  | 15 | 11 s | -3.47 | -5.13 | 1.48 | .002 | .003 |
|  |  |  | 15 | 12 s | -3.94 | -5.82 | 1.48 | < .001 | .001 |
|  |  |  | 15 | 13 s | -4.32 | -6.39 | 1.48 | < .001 | < .001 |
|  |  |  | 15 | 14 s | -4.67 | -6.90 | 1.48 | < .001 | < .001 |
|  |  |  | 15 | 15 s | -4.88 | -7.21 | 1.48 | < .001 | < .001 |
|  |  |  | 15 | 16 s | -5.00 | -7.38 | 1.48 | < .001 | < .001 |
|  |  |  | 15 | 17 s | -4.95 | -7.32 | 1.48 | < .001 | < .001 |
|  |  |  | 15 | 18 s | -4.88 | -7.21 | 1.48 | < .001 | < .001 |
|  |  |  | 15 | 19 s | -4.74 | -7.00 | 1.48 | < .001 | < .001 |
|  |  |  | 15 | 20 s | -4.72 | -6.97 | 1.48 | < .001 | < .001 |
| **LIFGop** | **16** | **3.36** | **15, 225** | **Overall time effect** |  |  |  | **< .001** | **< .001** |
|  |  |  | 15 | 5 s | -0.79 | -1.41 | 1.80 | .222 | .222 |
|  |  |  | 15 | 6 s | -0.98 | -1.75 | 1.80 | .172 | .184 |
|  |  |  | 15 | 7 s | -1.29 | -2.31 | 1.80 | .109 | .124 |
|  |  |  | 15 | 8 s | -1.58 | -2.84 | 1.80 | .068 | .083 |
|  |  |  | 15 | 9 s | -1.82 | -3.28 | 1.80 | .044 | .059 |
|  |  |  | 15 | 10 s | -2.15 | -3.86 | 1.80 | .024 | .035 |
|  |  |  | 15 | 11 s | -2.46 | -4.43 | 1.80 | .013 | .021 |
|  |  |  | 15 | 12 s | -2.78 | -5.00 | 1.80 | .007 | .012 |
|  |  |  | 15 | 13 s | -3.08 | -5.54 | 1.80 | .004 | .008 |
|  |  |  | 15 | 14 s | -3.50 | -6.29 | 1.80 | .002 | .004 |
|  |  |  | 15 | 15 s | -3.88 | -6.98 | 1.80 | < .001 | .002 |
|  |  |  | 15 | 16 s | -4.23 | -7.60 | 1.80 | < .001 | .001 |
|  |  |  | 15 | 17 s | -4.56 | -8.20 | 1.80 | < .001 | < .001 |
|  |  |  | 15 | 18 s | -4.80 | -8.63 | 1.80 | < .001 | < .001 |
|  |  |  | 15 | 19 s | -4.82 | -8.66 | 1.80 | < .001 | < .001 |
|  |  |  | 15 | 20 s | -4.74 | -8.53 | 1.80 | < .001 | < .001 |
| **LSTG** | **16** | **1.28** | **15, 225** | **Overall time effect** |  |  |  | **.217** | **.284** |
| **LMTG** | **16** | **1.15** | **15, 225** | **Overall time effect** |  |  |  | **.313** | **.355** |
| **LITG** | **15** | **1.55** | **15, 210** | **Overall time effect** |  |  |  | **.091** | **.155** |
| **LSMG** | **16** | **2.00** | **15, 225** | **Overall time effect** |  |  |  | **.017** | **.046** |
|  |  |  | 15 | 5 s | -1.38 | -1.91 | 1.38 | .093 | .093 |
|  |  |  | 15 | 6 s | -1.72 | -2.37 | 1.38 | .053 | .056 |
|  |  |  | 15 | 7 s | -2.01 | -2.77 | 1.38 | .032 | .036 |
|  |  |  | 15 | 8 s | -2.23 | -3.08 | 1.38 | .021 | .025 |
|  |  |  | 15 | 9 s | -2.35 | -3.24 | 1.38 | .017 | .022 |
|  |  |  | 15 | 10 s | -2.48 | -3.41 | 1.38 | .013 | .019 |
|  |  |  | 15 | 11 s | -2.60 | -3.58 | 1.38 | .010 | .016 |
|  |  |  | 15 | 12 s | -2.72 | -3.75 | 1.38 | .008 | .014 |
|  |  |  | 15 | 13 s | -2.97 | -4.09 | 1.38 | .005 | .010 |
|  |  |  | 15 | 14 s | -3.27 | -4.50 | 1.38 | .003 | .006 |
|  |  |  | 15 | 15 s | -3.53 | -4.86 | 1.38 | .002 | .004 |
|  |  |  | 15 | 16 s | -3.81 | -5.25 | 1.38 | < .001 | .003 |
|  |  |  | 15 | 17 s | -4.08 | -5.62 | 1.38 | < .001 | .002 |
|  |  |  | 15 | 18 s | -4.32 | -5.96 | 1.38 | < .001 | .002 |
|  |  |  | 15 | 19 s | -4.44 | -6.12 | 1.38 | < .001 | .002 |
|  |  |  | 15 | 20 s | -4.44 | -6.13 | 1.38 | < .001 | .002 |
| **LAG** | **14** | **1.11** | **15, 195** | **Overall time effect** |  |  |  | **.350** | **.371** |
| **RSFG** | **16** | **1.69** | **15, 225** | **Overall time effect** |  |  |  | **.054** | **.114** |
| **RMFG** | **16** | **2.35** | **15, 225** | **Overall time effect** |  |  |  | **.004** | **.013** |
|  |  |  | 15 | 5 s | 0.93 | 0.62 | 0.67 | .818 | .818 |
|  |  |  | 15 | 6 s | 0.89 | 0.59 | 0.67 | .805 | .818 |
|  |  |  | 15 | 7 s | 0.66 | 0.44 | 0.67 | .741 | .818 |
|  |  |  | 15 | 8 s | 0.39 | 0.26 | 0.67 | .649 | .799 |
|  |  |  | 15 | 9 s | 0.11 | 0.07 | 0.67 | .543 | .724 |
|  |  |  | 15 | 10 s | -0.30 | -0.20 | 0.67 | .384 | .558 |
|  |  |  | 15 | 11 s | -0.69 | -0.46 | 0.67 | .249 | .398 |
|  |  |  | 15 | 12 s | -1.03 | -0.68 | 0.67 | .160 | .285 |
|  |  |  | 15 | 13 s | -1.41 | -0.94 | 0.67 | .090 | .205 |
|  |  |  | 15 | 14 s | -1.71 | -1.14 | 0.67 | .054 | .177 |
|  |  |  | 15 | 15 s | -1.87 | -1.24 | 0.67 | .041 | .177 |
|  |  |  | 15 | 16 s | -1.96 | -1.31 | 0.67 | .034 | .177 |
|  |  |  | 15 | 17 s | -1.90 | -1.26 | 0.67 | .038 | .177 |
|  |  |  | 15 | 18 s | -1.69 | -1.13 | 0.67 | .055 | .177 |
|  |  |  | 15 | 19 s | -1.44 | -0.96 | 0.67 | .085 | .205 |
|  |  |  | 15 | 20 s | -1.11 | -0.74 | 0.67 | .142 | .283 |
| **RIFGtri** | **16** | **2.36** | **15, 225** | **Overall time effect** |  |  |  | **.004** | **.013** |
|  |  |  | 15 | 5 s | -0.39 | -0.52 | 1.31 | .350 | .350 |
|  |  |  | 15 | 6 s | -0.74 | -0.97 | 1.31 | .235 | .251 |
|  |  |  | 15 | 7 s | -1.20 | -1.58 | 1.31 | .124 | .142 |
|  |  |  | 15 | 8 s | -1.61 | -2.11 | 1.31 | .064 | .079 |
|  |  |  | 15 | 9 s | -1.99 | -2.62 | 1.31 | .032 | .043 |
|  |  |  | 15 | 10 s | -2.37 | -3.12 | 1.31 | .016 | .023 |
|  |  |  | 15 | 11 s | -2.66 | -3.49 | 1.31 | .009 | .014 |
|  |  |  | 15 | 12 s | -2.99 | -3.93 | 1.31 | .005 | .008 |
|  |  |  | 15 | 13 s | -3.29 | -4.33 | 1.31 | .002 | .005 |
|  |  |  | 15 | 14 s | -3.51 | -4.62 | 1.31 | .002 | .004 |
|  |  |  | 15 | 15 s | -3.69 | -4.85 | 1.31 | .001 | .003 |
|  |  |  | 15 | 16 s | -3.88 | -5.10 | 1.31 | < .001 | .002 |
|  |  |  | 15 | 17 s | -4.08 | -5.36 | 1.31 | < .001 | .002 |
|  |  |  | 15 | 18 s | -4.22 | -5.54 | 1.31 | < .001 | .002 |
|  |  |  | 15 | 19 s | -4.07 | -5.36 | 1.31 | < .001 | .002 |
|  |  |  | 15 | 20 s | -3.90 | -5.13 | 1.31 | < .001 | .002 |
| **RIFGop** | **16** | **1.65** | **15, 225** | **Overall time effect** |  |  |  | **.062** | **.117** |
| **RSTG** | **16** | **1.50** | **15, 225** | **Overall time effect** |  |  |  | **.106** | **.163** |
| **RMTG** | **16** | **1.96** | **15, 225** | **Overall time effect** |  |  |  | **.019** | **.046** |
|  |  |  | 15 | 5 s | -0.84 | -1.03 | 1.23 | .208 | .370 |
|  |  |  | 15 | 6 s | -0.73 | -0.90 | 1.23 | .240 | .383 |
|  |  |  | 15 | 7 s | -0.55 | -0.68 | 1.23 | .294 | .405 |
|  |  |  | 15 | 8 s | -0.45 | -0.56 | 1.23 | .329 | .405 |
|  |  |  | 15 | 9 s | -0.35 | -0.43 | 1.23 | .366 | .406 |
|  |  |  | 15 | 10 s | -0.26 | -0.32 | 1.23 | .399 | .406 |
|  |  |  | 15 | 11 s | -0.24 | -0.30 | 1.23 | .406 | .406 |
|  |  |  | 15 | 12 s | -0.49 | -0.61 | 1.23 | .314 | .405 |
|  |  |  | 15 | 13 s | -1.07 | -1.32 | 1.23 | .151 | .302 |
|  |  |  | 15 | 14 s | -1.60 | -1.97 | 1.23 | .065 | .149 |
|  |  |  | 15 | 15 s | -1.95 | -2.41 | 1.23 | .035 | .093 |
|  |  |  | 15 | 16 s | -2.15 | -2.65 | 1.23 | .024 | .077 |
|  |  |  | 15 | 17 s | -2.25 | -2.77 | 1.23 | .020 | .077 |
|  |  |  | 15 | 18 s | -2.30 | -2.83 | 1.23 | .018 | .077 |
|  |  |  | 15 | 19 s | -2.24 | -2.76 | 1.23 | .020 | .077 |
|  |  |  | 15 | 20 s | -2.16 | -2.66 | 1.23 | .024 | .077 |
| **RSMG** | **14** | **0.97** | **15, 195** | **Overall time effect** |  |  |  | **.489** | **.489** |
| **RAG** | **15** | **1.24** | **15, 210** | **Overall time effect** |  |  |  | **.244** | **.296** |

Note. Estimated marginal means (EMM) and standard error (SE) values are reported in µM. Model-level *q*-values reflect correction at the false discovery rate (FDR) across regions of interest (ROIs) within each Group x Task x Hemoglobin analysis set. EMM *q*-values reflect FDR correction across the 16 tested time points (5-20 s) within each ROI. Oxyhemoglobin (HbO) EMMs were tested against 0 in the positive direction; de-oxyhemoglobin (HbR) EMMs were tested against 0 in the negative direction. EMMs are shown only when the model-level FDR-corrected result is significant. AG = angular gyrus, IFGtri/op = inferior frontal gyrus, pars triangularis/opercularis, ITG = inferior temporal gyrus, L = left, MFG = middle frontal gyrus, MTG = middle temporal gyrus, R = right, SFG = superior frontal gyrus, SMG = supramarginal gyrus, STG = superior temporal gyrus.

Supplemental Table 6. Linear mixed effects models results for Rhyme Judgment for participants with aphasia

| **Region** | ***N*** | ***F*** | ***df*** | **Effect / time** | ***t*** | **EMM (µM)** | **SE (µM)** | ***p*** | ***q* (FDR)** |
| --- | --- | --- | --- | --- | --- | --- | --- | --- | --- |
| **HbO** |  |  |  |  |  |  |  |  |  |
| **LSFG** | **13** | **0.81** | **15, 180** | **Overall time effect** |  |  |  | **.671** | **.814** |
| **LMFG** | **13** | **0.32** | **15, 180** | **Overall time effect** |  |  |  | **.993** | **.993** |
| **LIFGtri** | **12** | **0.97** | **15, 165** | **Overall time effect** |  |  |  | **.486** | **.725** |
| **LIFGop** | **10** | **1.10** | **15, 135** | **Overall time effect** |  |  |  | **.359** | **.670** |
| **LSTG** | **6** | **1.76** | **15, 75** | **Overall time effect** |  |  |  | **.056** | **.160** |
| **LMTG** | **10** | **1.43** | **15, 135** | **Overall time effect** |  |  |  | **.141** | **.343** |
| **LITG** | **11** | **0.46** | **15, 150** | **Overall time effect** |  |  |  | **.957** | **.993** |
| **LSMG** | **11** | **3.01** | **15, 150** | **Overall time effect** |  |  |  | **< .001** | **.005** |
|  |  |  | 10 | 5 s | 0.32 | 1.68 | 5.27 | .378 | .378 |
|  |  |  | 10 | 6 s | 0.57 | 3.01 | 5.27 | .290 | .309 |
|  |  |  | 10 | 7 s | 0.83 | 4.36 | 5.27 | .213 | .244 |
|  |  |  | 10 | 8 s | 1.08 | 5.67 | 5.27 | .153 | .205 |
|  |  |  | 10 | 9 s | 1.37 | 7.23 | 5.27 | .100 | .160 |
|  |  |  | 10 | 10 s | 1.74 | 9.14 | 5.27 | .057 | .113 |
|  |  |  | 10 | 11 s | 2.22 | 11.70 | 5.27 | .025 | .067 |
|  |  |  | 10 | 12 s | 2.78 | 14.65 | 5.27 | .010 | .039 |
|  |  |  | 10 | 13 s | 3.07 | 16.19 | 5.27 | .006 | .039 |
|  |  |  | 10 | 14 s | 3.06 | 16.09 | 5.27 | .006 | .039 |
|  |  |  | 10 | 15 s | 2.86 | 15.09 | 5.27 | .008 | .039 |
|  |  |  | 10 | 16 s | 2.57 | 13.51 | 5.27 | .014 | .045 |
|  |  |  | 10 | 17 s | 2.08 | 10.94 | 5.27 | .032 | .074 |
|  |  |  | 10 | 18 s | 1.56 | 8.20 | 5.27 | .075 | .134 |
|  |  |  | 10 | 19 s | 1.09 | 5.75 | 5.27 | .150 | .205 |
|  |  |  | 10 | 20 s | 0.98 | 5.18 | 5.27 | .174 | .215 |
| **LAG** | **7** | **0.87** | **15, 90** | **Overall time effect** |  |  |  | **.594** | **.777** |
| **RSFG** | **13** | **0.95** | **15, 180** | **Overall time effect** |  |  |  | **.512** | **.725** |
| **RMFG** | **13** | **1.06** | **15, 180** | **Overall time effect** |  |  |  | **.394** | **.670** |
| **RIFGtri** | **13** | **0.53** | **15, 180** | **Overall time effect** |  |  |  | **.922** | **.993** |
| **RIFGop** | **13** | **1.20** | **15, 180** | **Overall time effect** |  |  |  | **.278** | **.591** |
| **RSTG** | **13** | **2.79** | **15, 180** | **Overall time effect** |  |  |  | **< .001** | **.005** |
|  |  |  | 12 | 5 s | 0.65 | 3.13 | 4.80 | .263 | .466 |
|  |  |  | 12 | 6 s | 0.57 | 2.72 | 4.80 | .291 | .466 |
|  |  |  | 12 | 7 s | 0.25 | 1.20 | 4.80 | .403 | .496 |
|  |  |  | 12 | 8 s | -0.09 | -0.41 | 4.80 | .533 | .569 |
|  |  |  | 12 | 9 s | -0.21 | -0.99 | 4.80 | .580 | .580 |
|  |  |  | 12 | 10 s | -0.04 | -0.21 | 4.80 | .517 | .569 |
|  |  |  | 12 | 11 s | 0.29 | 1.38 | 4.80 | .389 | .496 |
|  |  |  | 12 | 12 s | 0.72 | 3.45 | 4.80 | .243 | .466 |
|  |  |  | 12 | 13 s | 1.26 | 6.05 | 4.80 | .116 | .308 |
|  |  |  | 12 | 14 s | 1.79 | 8.59 | 4.80 | .049 | .201 |
|  |  |  | 12 | 15 s | 2.02 | 9.69 | 4.80 | .033 | .201 |
|  |  |  | 12 | 16 s | 1.96 | 9.40 | 4.80 | .037 | .201 |
|  |  |  | 12 | 17 s | 1.78 | 8.54 | 4.80 | .050 | .201 |
|  |  |  | 12 | 18 s | 1.41 | 6.77 | 4.80 | .092 | .294 |
|  |  |  | 12 | 19 s | 0.96 | 4.59 | 4.80 | .179 | .409 |
|  |  |  | 12 | 20 s | 0.47 | 2.25 | 4.80 | .324 | .471 |
| **RMTG** | **13** | **1.73** | **15, 180** | **Overall time effect** |  |  |  | **.049** | **.160** |
| **RSMG** | **13** | **1.93** | **15, 180** | **Overall time effect** |  |  |  | **.023** | **.097** |
| **RAG** | **13** | **2.27** | **15, 180** | **Overall time effect** |  |  |  | **.006** | **.033** |
|  |  |  | 12 | 5 s | 0.73 | 4.16 | 5.68 | .239 | .284 |
|  |  |  | 12 | 6 s | 0.89 | 5.08 | 5.68 | .194 | .283 |
|  |  |  | 12 | 7 s | 0.80 | 4.55 | 5.68 | .220 | .284 |
|  |  |  | 12 | 8 s | 0.64 | 3.65 | 5.68 | .266 | .284 |
|  |  |  | 12 | 9 s | 0.55 | 3.11 | 5.68 | .297 | .297 |
|  |  |  | 12 | 10 s | 0.68 | 3.85 | 5.68 | .256 | .284 |
|  |  |  | 12 | 11 s | 0.95 | 5.38 | 5.68 | .181 | .283 |
|  |  |  | 12 | 12 s | 1.29 | 7.32 | 5.68 | .111 | .222 |
|  |  |  | 12 | 13 s | 1.73 | 9.81 | 5.68 | .055 | .147 |
|  |  |  | 12 | 14 s | 2.23 | 12.66 | 5.68 | .023 | .091 |
|  |  |  | 12 | 15 s | 2.44 | 13.87 | 5.68 | .016 | .091 |
|  |  |  | 12 | 16 s | 2.42 | 13.78 | 5.68 | .016 | .091 |
|  |  |  | 12 | 17 s | 2.25 | 12.77 | 5.68 | .022 | .091 |
|  |  |  | 12 | 18 s | 1.90 | 10.79 | 5.68 | .041 | .131 |
|  |  |  | 12 | 19 s | 1.48 | 8.40 | 5.68 | .083 | .189 |
|  |  |  | 12 | 20 s | 1.11 | 6.32 | 5.68 | .144 | .256 |
| **HbR** |  |  |  |  |  |  |  |  |  |
| **LSFG** | **13** | **0.20** | **15, 180** | **Overall time effect** |  |  |  | **.999** | **1.000** |
| **LMFG** | **13** | **0.19** | **15, 180** | **Overall time effect** |  |  |  | **1.000** | **1.000** |
| **LIFGtri** | **12** | **1.41** | **15, 165** | **Overall time effect** |  |  |  | **.150** | **.510** |
| **LIFGop** | **10** | **0.46** | **15, 135** | **Overall time effect** |  |  |  | **.956** | **1.000** |
| **LSTG** | **6** | **1.89** | **15, 75** | **Overall time effect** |  |  |  | **.038** | **.216** |
| **LMTG** | **10** | **2.34** | **15, 135** | **Overall time effect** |  |  |  | **.005** | **.089** |
| **LITG** | **11** | **1.25** | **15, 150** | **Overall time effect** |  |  |  | **.242** | **.684** |
| **LSMG** | **11** | **0.71** | **15, 150** | **Overall time effect** |  |  |  | **.767** | **1.000** |
| **LAG** | **7** | **0.49** | **15, 90** | **Overall time effect** |  |  |  | **.941** | **1.000** |
| **RSFG** | **13** | **0.42** | **15, 180** | **Overall time effect** |  |  |  | **.972** | **1.000** |
| **RMFG** | **13** | **0.29** | **15, 180** | **Overall time effect** |  |  |  | **.996** | **1.000** |
| **RIFGtri** | **13** | **0.70** | **15, 180** | **Overall time effect** |  |  |  | **.782** | **1.000** |
| **RIFGop** | **13** | **1.19** | **15, 180** | **Overall time effect** |  |  |  | **.283** | **.687** |
| **RSTG** | **13** | **1.87** | **15, 180** | **Overall time effect** |  |  |  | **.029** | **.216** |
| **RMTG** | **13** | **1.03** | **15, 180** | **Overall time effect** |  |  |  | **.422** | **.798** |
| **RSMG** | **13** | **1.10** | **15, 180** | **Overall time effect** |  |  |  | **.357** | **.760** |
| **RAG** | **13** | **1.48** | **15, 180** | **Overall time effect** |  |  |  | **.116** | **.495** |

Note. Estimated marginal means (EMM) and standard error (SE) values are reported in µM. Model-level *q*-values reflect correction at the false discovery rate (FDR) across regions of interest (ROIs) within each Group x Task x Hemoglobin analysis set. EMM *q*-values reflect FDR correction across the 16 tested time points (5-20 s) within each ROI. Oxyhemoglobin (HbO) EMMs were tested against 0 in the positive direction; de-oxyhemoglobin (HbR) EMMs were tested against 0 in the negative direction. EMMs are shown only when the model-level FDR-corrected result is significant. AG = angular gyrus, IFGtri/op = inferior frontal gyrus, pars triangularis/opercularis, ITG = inferior temporal gyrus, L = left, MFG = middle frontal gyrus, MTG = middle temporal gyrus, R = right, SFG = superior frontal gyrus, SMG = supramarginal gyrus, STG = superior temporal gyrus.

Supplemental Table 7. Linear mixed effects models comparing Rhyme Judgment results between PWA and healthy controls

| **Region** | ***N* (PWA / Controls)** | ***F*** | ***df*** | **Effect** | ***t*** | **PWA - Controls (µM)** | | ***SE* (µM)** | ***p*** | ***q* (FDR)** |
| --- | --- | --- | --- | --- | --- | --- | --- | --- | --- | --- |
| **HbO** | | | | | | | | | | |
| LSFG | 13 / 16 | 0.03 | 1, 27 | Group |  | |  |  | .861 | .914 |
| LSFG | 13 / 16 | 1.06 | 15, 405 | Time |  | |  |  | .388 | .472 |
| LSFG | 13 / 16 | 1.02 | 15, 405 | Group x Time |  | |  |  | .435 | .873 |
| LMFG | 13 / 16 | 0.00 | 1, 27 | Group |  | |  |  | .950 | .950 |
| LMFG | 13 / 16 | 0.53 | 15, 405 | Time |  | |  |  | .927 | .970 |
| LMFG | 13 / 16 | 0.71 | 15, 405 | Group x Time |  | |  |  | .771 | .873 |
| LIFGtri | 12 / 16 | 0.27 | 1, 26 | Group |  | |  |  | .608 | .783 |
| LIFGtri | 12 / 16 | 1.36 | 15, 390 | Time |  | |  |  | .166 | .283 |
| LIFGtri | 12 / 16 | 0.80 | 15, 390 | Group x Time |  | |  |  | .683 | .873 |
| LIFGop | 10 / 16 | 0.35 | 1, 24 | Group |  | |  |  | .560 | .783 |
| LIFGop | 10 / 16 | 1.62 | 15, 360 | Time |  | |  |  | .065 | .159 |
| LIFGop | 10 / 16 | 0.56 | 15, 360 | Group x Time |  | |  |  | .906 | .906 |
| LSTG | 6 / 16 | 0.46 | 1, 20 | Group |  | |  |  | .508 | .783 |
| LSTG | 6 / 16 | 1.15 | 15, 300 | Time |  | |  |  | .309 | .437 |
| LSTG | 6 / 16 | 0.74 | 15, 300 | Group x Time |  | |  |  | .742 | .873 |
| LMTG | 10 / 16 | 1.28 | 1, 24 | Group |  | |  |  | .270 | .783 |
| LMTG | 10 / 16 | 1.07 | 15, 360 | Time |  | |  |  | .387 | .472 |
| LMTG | 10 / 16 | 0.90 | 15, 360 | Group x Time |  | |  |  | .564 | .873 |
| LITG | 11 / 15 | 2.30 | 1, 24 | Group |  | |  |  | .143 | .783 |
| LITG | 11 / 15 | 0.43 | 15, 360 | Time |  | |  |  | .970 | .970 |
| LITG | 11 / 15 | 0.94 | 15, 360 | Group x Time |  | |  |  | .515 | .873 |
| LSMG | 11 / 16 | 0.31 | 1, 25 | Group |  | |  |  | .582 | .783 |
| LSMG | 11 / 16 | 4.39 | 15, 375 | Time |  | |  |  | < .001 | < .001 |
| LSMG | 11 / 16 | 1.41 | 15, 375 | Group x Time |  | |  |  | .141 | .601 |
| LAG | 7 / 14 | 0.11 | 1, 19 | Group |  | |  |  | .743 | .843 |
| LAG | 7 / 14 | 1.43 | 15, 285 | Time |  | |  |  | .131 | .247 |
| LAG | 7 / 14 | 0.62 | 15, 285 | Group x Time |  | |  |  | .858 | .906 |
| RSFG | 13 / 16 | 0.24 | 1, 27 | Group |  | |  |  | .625 | .783 |
| RSFG | 13 / 16 | 1.27 | 15, 405 | Time |  | |  |  | .218 | .336 |
| RSFG | 13 / 16 | 0.88 | 15, 405 | Group x Time |  | |  |  | .591 | .873 |
| RMFG | 13 / 16 | 0.26 | 1, 27 | Group |  | |  |  | .617 | .783 |
| RMFG | 13 / 16 | 1.44 | 15, 405 | Time |  | |  |  | .126 | .247 |
| RMFG | 13 / 16 | 1.00 | 15, 405 | Group x Time |  | |  |  | .457 | .873 |
| RIFGtri | 13 / 16 | 0.53 | 1, 27 | Group |  | |  |  | .471 | .783 |
| RIFGtri | 13 / 16 | 0.72 | 15, 405 | Time |  | |  |  | .762 | .863 |
| RIFGtri | 13 / 16 | 0.82 | 15, 405 | Group x Time |  | |  |  | .658 | .873 |
| RIFGop | 13 / 16 | 0.26 | 1, 27 | Group |  | |  |  | .612 | .783 |
| RIFGop | 13 / 16 | 1.85 | 15, 405 | Time |  | |  |  | .026 | .074 |
| RIFGop | 13 / 16 | 0.97 | 15, 405 | Group x Time |  | |  |  | .481 | .873 |
| RSTG | 13 / 16 | 0.82 | 1, 27 | Group |  | |  |  | .374 | .783 |
| RSTG | 13 / 16 | 4.27 | 15, 405 | Time |  | |  |  | < .001 | < .001 |
| RSTG | 13 / 16 | 2.70 | 15, 405 | Group x Time |  | |  |  | < .001 | .010 |
|  |  |  | 27 | 5 s | 0.90 | | 4.54 | 5.02 | .374 | .598 |
|  |  |  | 27 | 6 s | 0.94 | | 4.71 | 5.02 | .356 | .598 |
|  |  |  | 27 | 7 s | 0.64 | | 3.23 | 5.02 | .526 | .765 |
|  |  |  | 27 | 8 s | 0.21 | | 1.07 | 5.02 | .832 | .937 |
|  |  |  | 27 | 9 s | -0.08 | | -0.40 | 5.02 | .937 | .937 |
|  |  |  | 27 | 10 s | -0.10 | | -0.52 | 5.02 | .918 | .937 |
|  |  |  | 27 | 11 s | 0.08 | | 0.41 | 5.02 | .935 | .937 |
|  |  |  | 27 | 12 s | 0.44 | | 2.23 | 5.02 | .660 | .880 |
|  |  |  | 27 | 13 s | 1.05 | | 5.29 | 5.02 | .302 | .598 |
|  |  |  | 27 | 14 s | 1.76 | | 8.82 | 5.02 | .090 | .289 |
|  |  |  | 27 | 15 s | 2.20 | | 11.02 | 5.02 | .037 | .197 |
|  |  |  | 27 | 16 s | 2.28 | | 11.47 | 5.02 | .030 | .197 |
|  |  |  | 27 | 17 s | 2.23 | | 11.17 | 5.02 | .035 | .197 |
|  |  |  | 27 | 18 s | 2.01 | | 10.08 | 5.02 | .055 | .220 |
|  |  |  | 27 | 19 s | 1.66 | | 8.33 | 5.02 | .109 | .289 |
|  |  |  | 27 | 20 s | 1.32 | | 6.65 | 5.02 | .197 | .450 |
| RMTG | 13 / 16 | 0.22 | 1, 27 | Group |  | |  |  | .645 | .783 |
| RMTG | 13 / 16 | 2.23 | 15, 405 | Time |  | |  |  | .005 | .018 |
| RMTG | 13 / 16 | 2.22 | 15, 405 | Group x Time |  | |  |  | .006 | .047 |
|  |  |  | 27 | 5 s | 0.47 | | 3.05 | 6.54 | .645 | .935 |
|  |  |  | 27 | 6 s | 0.41 | | 2.66 | 6.54 | .687 | .935 |
|  |  |  | 27 | 7 s | 0.11 | | 0.71 | 6.54 | .914 | .936 |
|  |  |  | 27 | 8 s | -0.24 | | -1.58 | 6.54 | .810 | .935 |
|  |  |  | 27 | 9 s | -0.47 | | -3.05 | 6.54 | .644 | .935 |
|  |  |  | 27 | 10 s | -0.45 | | -2.97 | 6.54 | .653 | .935 |
|  |  |  | 27 | 11 s | -0.35 | | -2.30 | 6.54 | .728 | .935 |
|  |  |  | 27 | 12 s | -0.23 | | -1.52 | 6.54 | .818 | .935 |
|  |  |  | 27 | 13 s | 0.08 | | 0.53 | 6.54 | .936 | .936 |
|  |  |  | 27 | 14 s | 0.57 | | 3.74 | 6.54 | .572 | .935 |
|  |  |  | 27 | 15 s | 0.88 | | 5.76 | 6.54 | .386 | .935 |
|  |  |  | 27 | 16 s | 1.02 | | 6.64 | 6.54 | .319 | .935 |
|  |  |  | 27 | 17 s | 1.15 | | 7.50 | 6.54 | .261 | .935 |
|  |  |  | 27 | 18 s | 1.18 | | 7.73 | 6.54 | .247 | .935 |
|  |  |  | 27 | 19 s | 1.16 | | 7.55 | 6.54 | .258 | .935 |
|  |  |  | 27 | 20 s | 1.09 | | 7.14 | 6.54 | .284 | .935 |
| RSMG | 13 / 14 | 0.39 | 1, 25 | Group |  | |  |  | .537 | .783 |
| RSMG | 13 / 14 | 2.47 | 15, 375 | Time |  | |  |  | .002 | .008 |
| RSMG | 13 / 14 | 1.07 | 15, 375 | Group x Time |  | |  |  | .380 | .873 |
| RAG | 13 / 15 | 0.42 | 1, 26 | Group |  | |  |  | .524 | .783 |
| RAG | 13 / 15 | 3.07 | 15, 390 | Time |  | |  |  | < .001 | < .001 |
| RAG | 13 / 15 | 1.49 | 15, 390 | Group x Time |  | |  |  | .107 | .601 |
| **HbR** | | | | | | | | | | |
| LSFG | 13 / 16 | 0.00 | 1, 27 | Group |  | |  |  | .970 | .975 |
| LSFG | 13 / 16 | 0.21 | 15, 405 | Time |  | |  |  | .999 | .999 |
| LSFG | 13 / 16 | 0.64 | 15, 405 | Group x Time |  | |  |  | .842 | .928 |
| LMFG | 13 / 16 | 0.15 | 1, 27 | Group |  | |  |  | .701 | .975 |
| LMFG | 13 / 16 | 0.25 | 15, 405 | Time |  | |  |  | .998 | .999 |
| LMFG | 13 / 16 | 0.62 | 15, 405 | Group x Time |  | |  |  | .861 | .928 |
| LIFGtri | 12 / 16 | 0.45 | 1, 26 | Group |  | |  |  | .511 | .975 |
| LIFGtri | 12 / 16 | 0.87 | 15, 390 | Time |  | |  |  | .597 | .923 |
| LIFGtri | 12 / 16 | 1.62 | 15, 390 | Group x Time |  | |  |  | .066 | .375 |
| LIFGop | 10 / 16 | 0.00 | 1, 24 | Group |  | |  |  | .975 | .975 |
| LIFGop | 10 / 16 | 0.46 | 15, 360 | Time |  | |  |  | .961 | .999 |
| LIFGop | 10 / 16 | 0.90 | 15, 360 | Group x Time |  | |  |  | .569 | .880 |
| LSTG | 6 / 16 | 0.03 | 1, 20 | Group |  | |  |  | .857 | .975 |
| LSTG | 6 / 16 | 1.55 | 15, 300 | Time |  | |  |  | .087 | .370 |
| LSTG | 6 / 16 | 0.55 | 15, 300 | Group x Time |  | |  |  | .913 | .928 |
| LMTG | 10 / 16 | 0.54 | 1, 24 | Group |  | |  |  | .469 | .975 |
| LMTG | 10 / 16 | 2.46 | 15, 360 | Time |  | |  |  | .002 | .032 |
| LMTG | 10 / 16 | 0.52 | 15, 360 | Group x Time |  | |  |  | .928 | .928 |
| LITG | 11 / 15 | 0.23 | 1, 24 | Group |  | |  |  | .634 | .975 |
| LITG | 11 / 15 | 1.26 | 15, 360 | Time |  | |  |  | .225 | .611 |
| LITG | 11 / 15 | 0.61 | 15, 360 | Group x Time |  | |  |  | .865 | .928 |
| LSMG | 11 / 16 | 0.00 | 1, 25 | Group |  | |  |  | .971 | .975 |
| LSMG | 11 / 16 | 1.13 | 15, 375 | Time |  | |  |  | .324 | .611 |
| LSMG | 11 / 16 | 0.99 | 15, 375 | Group x Time |  | |  |  | .463 | .787 |
| LAG | 7 / 14 | 0.76 | 1, 19 | Group |  | |  |  | .394 | .975 |
| LAG | 7 / 14 | 1.17 | 15, 285 | Time |  | |  |  | .297 | .611 |
| LAG | 7 / 14 | 1.24 | 15, 285 | Group x Time |  | |  |  | .244 | .691 |
| RSFG | 13 / 16 | 1.09 | 1, 27 | Group |  | |  |  | .307 | .975 |
| RSFG | 13 / 16 | 0.51 | 15, 405 | Time |  | |  |  | .934 | .999 |
| RSFG | 13 / 16 | 1.40 | 15, 405 | Group x Time |  | |  |  | .141 | .518 |
| RMFG | 13 / 16 | 0.15 | 1, 27 | Group |  | |  |  | .703 | .975 |
| RMFG | 13 / 16 | 0.42 | 15, 405 | Time |  | |  |  | .972 | .999 |
| RMFG | 13 / 16 | 0.73 | 15, 405 | Group x Time |  | |  |  | .750 | .928 |
| RIFGtri | 13 / 16 | 0.04 | 1, 27 | Group |  | |  |  | .852 | .975 |
| RIFGtri | 13 / 16 | 0.64 | 15, 405 | Time |  | |  |  | .844 | .999 |
| RIFGtri | 13 / 16 | 1.72 | 15, 405 | Group x Time |  | |  |  | .045 | .375 |
| RIFGop | 13 / 16 | 0.13 | 1, 27 | Group |  | |  |  | .719 | .975 |
| RIFGop | 13 / 16 | 1.18 | 15, 405 | Time |  | |  |  | .285 | .611 |
| RIFGop | 13 / 16 | 1.38 | 15, 405 | Group x Time |  | |  |  | .152 | .518 |
| RSTG | 13 / 16 | 0.71 | 1, 27 | Group |  | |  |  | .406 | .975 |
| RSTG | 13 / 16 | 1.25 | 15, 405 | Time |  | |  |  | .229 | .611 |
| RSTG | 13 / 16 | 1.06 | 15, 405 | Group x Time |  | |  |  | .393 | .742 |
| RMTG | 13 / 16 | 0.84 | 1, 27 | Group |  | |  |  | .366 | .975 |
| RMTG | 13 / 16 | 1.56 | 15, 405 | Time |  | |  |  | .083 | .370 |
| RMTG | 13 / 16 | 2.05 | 15, 405 | Group x Time |  | |  |  | .012 | .196 |
| RSMG | 13 / 14 | 0.11 | 1, 25 | Group |  | |  |  | .741 | .975 |
| RSMG | 13 / 14 | 0.97 | 15, 375 | Time |  | |  |  | .483 | .821 |
| RSMG | 13 / 14 | 1.13 | 15, 375 | Group x Time |  | |  |  | .329 | .742 |
| RAG | 13 / 15 | 1.65 | 1, 26 | Group |  | |  |  | .210 | .975 |
| RAG | 13 / 15 | 1.74 | 15, 390 | Time |  | |  |  | .041 | .352 |
| RAG | 13 / 15 | 1.09 | 15, 390 | Group x Time |  | |  |  | .361 | .742 |

Note. Differences between PWA and controls in estimated marginal means and standard error (SE) values are reported in µM. Model-level *q*-values reflect correction at the false discovery rate (FDR) across regions of interest (ROIs) within each Group x Task x Hemoglobin analysis set. EMM *q*-values reflect FDR correction across the 16 tested time points (5-20 s) within each ROI. Oxyhemoglobin (HbO) EMMs were tested against 0 in the positive direction; de-oxyhemoglobin (HbR) EMMs were tested against 0 in the negative direction. EMMs are shown only when the model-level FDR-corrected result for the Group x Time interaction is significant. AG = angular gyrus, IFGtri/op = inferior frontal gyrus, pars triangularis/opercularis, ITG = inferior temporal gyrus, L = left, MFG = middle frontal gyrus, MTG = middle temporal gyrus, R = right, SFG = superior frontal gyrus, SMG = supramarginal gyrus, STG = superior temporal gyrus.

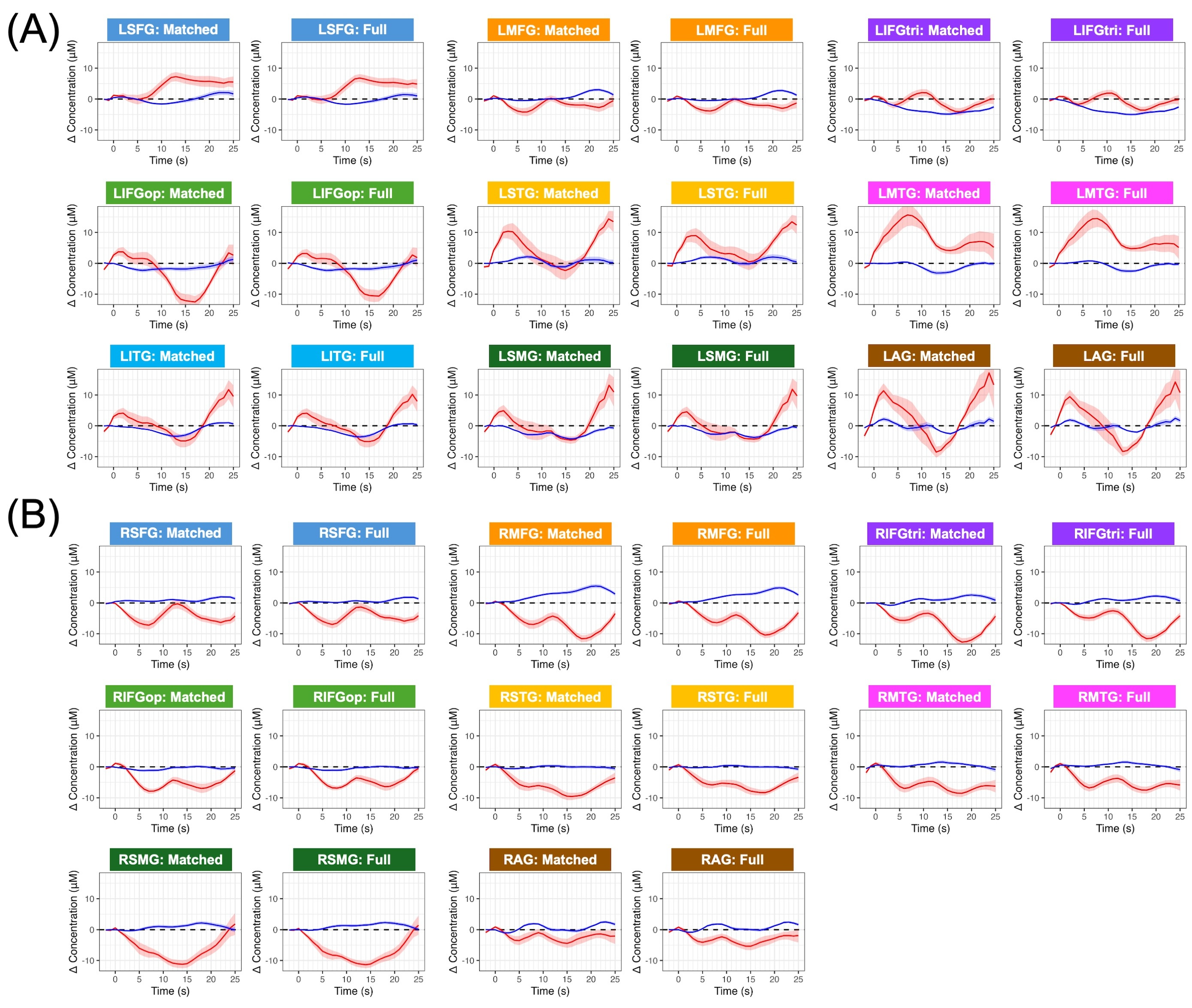

Supplemental Figure 1. Changes in oxyhemoglobin (in red) and deoxyhemoglobin (in blue) for left hemisphere (A) and right hemisphere (B) regions of interest during Semantic Matching. The left-sided plot for each region shows the group-averaged hemodynamic GLM output for the 13 participants with aphasia who completed both Semantic Matching and Rhyme Judgment. The right-sided plot for each region shows the group-averaged hemodynamic GLM output for the full sample of 15 participants with aphasia who completed the Semantic Matching task. Shaded ribbons represent within-group standard error around the mean time series. AG = angular gyrus, IFGtri/op = inferior frontal gyrus, pars triangularis/opercularis, ITG = inferior temporal gyrus, L = left, MFG = middle frontal gyrus, MTG = middle temporal gyrus, R = right, SFG = superior frontal gyrus, SMG = supramarginal gyrus, STG = superior temporal gyrus.
